# Correlates of circulating extracellular vesicle cargo with key clinical features of type 1 diabetes

**DOI:** 10.1101/2022.03.10.22272207

**Authors:** Anna Casu, Yury O. Nunez Lopez, Gongxin Yu, Christopher Clifford, Anika Bilal, Alejandra M. Petrilli, Heather Cornnell, Karen Corbin, Anton Iliuk, David Maahs, Elizabeth J. Mayer-Davis, Richard E. Pratley

## Abstract

Type 1 diabetes (T1D) is a heterogeneous disease with a slower evolution in individuals diagnosed at older ages. There are no validated clinical or laboratory biomarkers to predict the rate of insulin secretion decline either before or after the clinical onset of the disease, or the rate of progression to chronic complications of the disease. This pilot study aimed to characterize the proteomic and phosphoproteomic landscape of circulating extracellular vesicles (EVs) across a range of obesity in carefully matched established T1D and control subjects. We used archived serum samples from 17 human subjects (N=10 with T1D and N=7 normal healthy volunteers) from the ACME study (NCT03379792). EVs were isolated using EVtrap® technology (Tymora). Mass spectrometry-based methods were used to detect the global circulating EV proteome and phosphoproteome. Differential expression, coexpression network (WGCNA), and pathway enrichment analyses were implemented. The detected proteins and phosphoproteins were highly enriched (75%) in exosomal proteins cataloged in the ExoCarta database. A total of 181 differentially expressed EV proteins and 15 differentially expressed EV phosphoproteins were identified, with 8 upregulated EV proteins (i.e., CD63, RAB14, VCP, BSG, FLNA, GNAI2, LAMP2, and EZR) and 1 downregulated EV phosphoprotein (i.e., TUBA1B) listed among the top 100 ExoCarta proteins. This suggests that T1D could indeed modulate EV biogenesis and secretion. Enrichment analyses of both differentially expressed EV proteins and EV phosphoproteins identified associations of upregulated features with neutrophil, platelet, and immune response functions, as well as prion disease and other neurodegenerative diseases, among others. On the other hand, downregulated EV proteins were involved in MHC class II signaling and the regulation of monocyte differentiation. Potential novel key roles in T1D for C1q, plasminogen, IL6ST, CD40, HLA-DQB1, and phosphorylated S100A9, are highlighted. Remarkably, WGCNA uncovered two protein modules significantly associated with pancreas size, which may be implicated in the pathogenesis of T1D. Similarly, these modules showed significant enrichment for membrane compartments, processes associated with inflammation and the immune response, and regulation of viral processes, among others. This study demonstrates the potential of EV proteomic and phosphoproteomic signatures to provide insight into the pathobiology of type 1 diabetes and its complications.

## Introduction

Type 1 diabetes (T1D) is a chronic disease characterized by hyperglycemia due to dysfunction and loss of β-cells and the consequent lack of endogenous insulin secretion. The β-cell loss of function and mass is due to a targeted autoimmune process against these cells, but the precise cause and pathological mechanisms are still largely unknown. Even if this description the pathogenic mechanism and evolution seem straightforward, the disease is heterogeneous with a slower evolution in individuals diagnosed at older ages (>18 years of age) and there are no clinical or laboratory biomarkers means to predict the rate of insulin secretion decline either before or after the clinical onset of the disease (1-3).

Once established, T1D is treated with subcutaneous insulin substitution to achieve blood and tissue glucose levels sufficiently low to prevent or limit the onset of micro and macrovascular chronic complications of the disease (4). However, there is a heterogeneity in the progression to chronic complications that is not completely explained by glycemic control. This is particularly relevant when considering the risk of cardiovascular disease in patients with T1D (5). The cardiovascular risk should be interpreted in the context of the obesity epidemic that does not spare the T1D population and it is possibly exacerbated by insulin treatment itself (6). Additional biomarkers are therefore needed to improve the diagnostic capacity, predict progression and risk of chronic complications, and evaluate treatment efficacy.

In recent years, attention have been directed to the rapidly evolving research area addressing the role of extracellular vesicles (EVs) as vehicles of intercellular communications. EVs in their different forms (i.e., exosomes and microvesicles) originating from virtually all tissues can be found in the peripheral circulation and have been studied as non-invasive disease biomarkers, particularly in oncology (7). They can contribute to the activation and regulation of physiologic and pathologic responses to β-cell stress or may contribute to the activation of the autoimmune process. On the other hand, they may serve as biomarkers of metabolic derangement, insulin resistance, and chronic complications in diabetes (8).

The use of the EVs as a biomarker in T1D requires a proper understanding of the effect of different clinical components of the disease on the characteristics of circulating EVs, particularly on the EV proteome. The interest in the EV proteome resides in the possibility of identifying disease-specific signatures that, even if not directly pathogenetic, may be used as predictors of specific phenotypes or disease progression.

In this cross-sectional pilot study, we describe the proteome of serum-derived EVs obtained from individuals with T1D who have been deeply metabolically phenotyped and compare it with matched healthy controls. Our ultimate goal was the identification of biomarkers that may improve our understanding of overweight and obesity in T1D. In addition, to gain insight into EV proteomic signatures associated with either pathogenesis-specific or hyperglycemia-related clinical characteristics, we implemented a novel approach based on weighted gene coexpression network analysis (WGCNA) of the EV proteome and an extensive list of clinical metabolic features.

## Materials and Methods

### Study population

The study was conducted according to the principles of the Declaration of Helsinki and followed GCP guidelines. All procedures were approved by the AdventHealth Translational Research Institute Institutional (AH/TRI) Review Board. Informed consent was obtained from all volunteers before initiation of the study. The goal of this pilot study was to characterize EVs across a range of obesity in carefully matched established T1D and Control groups. Archived serum samples from 17 human subjects (N=10 with T1D and N=7 healthy volunteers with normal glucose tolerance (NGT) from the ACME study (NCT03379792) were used. Participants were specifically selected as a subgroup that was well balanced for most characteristics. This selection was automated using custom script based on the *MatchIt* package (9) in the R programing environment.

### In-depth clinical and metabolic phenotype

Anthropometric measures were performed according to standardized protocols. Fasting blood samples were obtained. Plasma glucose concentrations were measured using the glucose oxidase method with a YSI 2300 STAT Plus Analyzer (YSI Life Sciences, Yellow Springs, OH). HbA1c levels were measured using a Cobas Integra 800 Analyzer (Roche, Basel, Switzerland). Body composition was measured using a GE Lunar iDEXA whole-body scanner (GE, Madison, WI). Whole body magnetic resonance imaging (MRI) was implemented using a Philips 3T Achieva MRI instrument (Philips Medical Systems MR Inc., Latham, NY). Metabolic assessment in whole room calorimeters at AH/TRI according to standard methodology (10) allowed the simultaneous measure of total energy expenditure and substrate oxidation in a free-living environment with multiple activities. Continuous glucose monitoring (CGM) Dexcom G4 (Dexcom Inc., San Diego, CA, USA) was used to monitor glucose levels that were summarized according to what was proposed by Battelino et al. (11). A list of 154 variables included in the study is presented in Supplementary Table ST1.

### EV isolation using Tymora’s EVtrap technology

Blood samples were allowed to coagulate at room temperature for 30 minutes after collection, then centrifuged at 1500 ×g for 15 min at 4 °C to produce serum. Serum was stored at -80 °C until use. Frozen serum samples were thawed, then spun at 2,500 × g for 10 minutes. The pre-cleared samples were then diluted 20-fold in PBS and incubated with EVtrap beads for 30 min (12). After supernatant removal using a magnetic separator rack, the beads were washed with PBS, and the EVs were eluted by a 10 min incubation with 200 mM triethylamine (TEA, Millipore-Sigma) and the resulting EV samples fully dried in a vacuum centrifuge.

### Nanoparticle Tracking Analysis (NTA)

The size distribution and concentration of particles in EV preparations were analyzed using dynamic light-scattering technology with a NanoSight NS300 instrument and NTA-3.4 software (Malvern Panalytical, Malvern). The instrument was equipped with a 488 nm blue laser module, flow-cell top plate, integrated temperature control, and a single-syringe pump module. Samples were diluted using cell culture grade water (Corning cat# 25-005-CI) to produce an optimal particle concentration for final measurement in the range of 10^7^ to 10^9^ particles/ml. Final quantification included 5 standard measurements of 1 minute of duration each, taken at a controlled temperature of 25°C and under constant automatic flow. Camera level for video capture was set to 12 and detection threshold to 5 for all sample measurements.

### Mass spectrometry (LC-MS/MS)-based methods used to detect the global EV proteome and phosphoproteome

The isolated and dried EV samples were processed as described previously (https://pubmed.ncbi.nlm.nih.gov/32396726/). Briefly, EV samples were lysed to extract proteins using the phase-transfer surfactant (PTS) aided procedure (13) and the proteins digested with Lys-C (Wako) at 1:100 (wt/wt) enzyme-to-protein ratio for 3 h at 37°C. Trypsin was added to a final 1:50 (wt/wt) enzyme-to-protein ratio for overnight digestion at 37°C. After surfactant removal, the resulting peptides were desalted using Top-Tip C18 tips (Glygen) according to manufacturer’s instructions. Each sample was split into 99% and 1% aliquots for phosphoproteomic and proteomic experiments respectively. The samples were dried completely in a vacuum centrifuge and stored at -80°C. For phosphoproteome analysis, the 99% portion of each sample was subjected to phosphopeptide enrichment using PolyMAC Phosphopeptide Enrichment kit (Tymora Analytical) according to manufacturer’s instructions, and the eluted phosphopeptides dried completely in a vacuum centrifuge. For phosphoproteomics analysis the whole enriched sample was used, while for proteomics only 50% of the sample was loaded onto the LC-MS.

Each dried peptide or phosphopeptide sample was dissolved at 0.1 μg/μL in 0.05% trifluoroacetic acid with 3% (vol/vol) acetonitrile. 10 μL of each sample was injected into an Ultimate 3000 nano UHPLC system (Thermo Fisher Scientific). Peptides were captured on a 2-cm Acclaim PepMap trap column and separated on a heated 50-cm column packed with ReproSil Saphir 1.9 μm C18 beads (Dr. Maisch GmbH). The mobile phase buffer consisted of 0.1% formic acid in ultrapure water (buffer A) with an eluting buffer of 0.1% formic acid in 80% (vol/vol) acetonitrile (buffer B) run with a linear 60-min gradient of 6–30% buffer B at flow rate of 300 nL/min. The UHPLC was coupled online with a Q-Exactive HF-X mass spectrometer (Thermo Fisher Scientific). The mass spectrometer was operated in the data-dependent mode, in which a full-scan MS (from m/z 375 to 1,500 with the resolution of 60,000) was followed by MS/MS of the 15 most intense ions (30,000 resolution; normalized collision energy - 28%; automatic gain control target (AGC) - 2E4, maximum injection time - 200 ms; 60sec exclusion].

### Bioinformatic analysis

The raw files were searched directly against the human Uniprot database with no redundant entries, using Byonic (Protein Metrics) and Sequest search engines loaded into Proteome Discoverer 2.3 software (Thermo Fisher Scientific). MS1 precursor mass tolerance was set at 10 ppm, and MS2 tolerance was set at 20ppm. Search criteria included a static carbamidomethylation of cysteines (+57.0214 Da), and variable modifications of oxidation (+15.9949 Da) on methionine residues, acetylation (+42.011 Da) at N terminus of proteins, and phosphorylation of S, T and Y residues (+79.996 Da) for the phosphoproteomics data. Search was performed with full trypsin/P digestion and allowed a maximum of two missed cleavages on the peptides analyzed from the sequence database. The false-discovery rates of proteins and peptides were set at 0.01. All protein and peptide identifications were grouped and any redundant entries were removed. Only unique peptides and unique master proteins were reported.

All data were quantified using the label-free quantitation node of Precursor Ions Quantifier through the Proteome Discoverer v2.3 (Thermo Fisher Scientific). For the quantification of proteomic or phosphoproteomic data, the intensities of peptides/phosphopeptides were extracted with initial precursor mass tolerance set at 10 ppm, minimum number of isotope peaks as 2, maximum ΔRT of isotope pattern multiplets – 0.2 min, PSM confidence FDR of 0.01, with hypothesis test of ANOVA, maximum RT shift of 5 min, pairwise ratio-based ratio calculation, and 100 as the maximum allowed fold change. The abundance levels of all peptides and proteins were normalized using the total peptide amount normalization node in the Proteome Discoverer. For calculations of fold-change between the groups of proteins, total protein abundance values were added together and the ratios of these sums were used to compare proteins within different samples.

The EV-proteomic and phosphoproteomic expression profiles were analyzed with a user-defined bioinformatic procedure that included raw data preprocessing, differential expression analysis, weighted-gene correlation network analysis (WGCNA), and enriched functional analyses. Briefly, missing values in the expression profiles were imputed, and the data were then log2-transformed, and scale-standardized. The imputation was performed with random forest, an advanced machine-learning algorithm (14) known to be adaptive to interactions and nonlinearity with capability to handle mixed types of missing data (15). The normalized data were then analyzed using pvca (16) and limma (17), two R/Bioconductor software packages for biomarker discovery. Specifically, pvca, a package for principal variance component analysis, was first applied to identify significant covariates by fitting all “sources” as random effects, then linear models were created incorporating those significant covariates. Once established, the linear models were fitted using weighted least squares for each protein, moderated t-statistics, moderated F-statistic, log-fold changes and p-values of differential expression were calculated by empirical Bayes moderation of the standard errors. Finally, the Benjamin-Hochberg (BH) method was used to adjust the p-values. The network module analysis was performed with WGCNA, an R package for weighted gene correlation network analysis (18). In short, it first performs a weighted protein co-expression network analysis to find clusters of highly correlated proteins (modules), and then relates modules to the clinical measurements. Subsequent module membership, gene-trait significance and intra-module connectivity analysis was applied to identify the key driver proteins in modules of interest. KEGG pathway and Gene Ontology enrichment analyses were performed on the sets of differentially expressed proteins and phosphoproteins with clusterProfiler, an R package for enriched function analysis (19). Given a set of highly significant proteins or phosphoproteins identified, clusterProfiler suggests KEGG pathways and GO Ontology functional groups significantly affected.

### Statistical analysis

Data normality was tested using the Shapiro-Wilk test, and nonnormal data was log-transformed to approximate normality. Differences in baseline clinical characteristics were assessed using the Welch two-sample t test (for continuous variables) or the Fisher exact test (for categorical variables). For assessment of differential expression in EV proteins and phosphoproteins, linear models using the *limma* package were implemented. Linear models included sex as covariate. Calculated effects and correlations with two-tailed P<0.05 and FDR<0.1 (for proteomics) or FDR<0.2 (for phosphoproteomics) were considered significant. False discovery rates (FDR) correcting for multiple testing were calculated using the Benjamini-Hochberg correction as implemented for the *p*.*adjust* function in the *stats* package.

## Results

### Study design and clinical characteristics of the study cohort

A balanced subset of 17 participants from the ACME study was selected for this pilot proteomic and phosphoproteomic study. The subset was not confounded by differences in variables known to affect metabolic function (e.g., age, BMI, among others). Archived sera from these participants were available for EV purification and proteome/phosphoproteome analysis. The average age of the participants was 25.7 years (range 20.1-34.5), 11/17 were females, and the average BMI was 26.8 kg/m2 (range 19.5-38.6) for the whole population. The comparison of 154 anthropometric, clinical, and metabolic characteristics for the study cohort are presented in Supplementary Table ST1.

### EV purification and characterization demonstrate enrichment in exosomal particles

The EVtrap efficiently isolated circulating particles with a distribution of sizes compatible with exosomes (Figure 1A). There was a trend (p = 0.07) to higher modal particle sizes in the T1D group (the mode of particle size was 151.5 nm in T1D compared to 137.7 nm in controls). However, the average particle size was comparable between the groups (mean size 209.5 nm in T1D and 201.3 nm in controls, p = 0.159). Similarly, the average particle concentration did not differ between the two groups (3.6×10^10^ particles/ml in T1D and 4.6×10^10^ particles/mL in controls, p = 0.346) (Figure 1A). LC-MS/MS of isolated EVs from participant sera identified 1950 EV proteins in 1 ml of serum, which included 1467 (75.2%) proteins reported in ExoCarta (a database collecting information on all exosomal proteins, RNA, and lipids (20)), and more specifically, 91 (91%) of the top 100 exosomal proteins reported in this database (Figure 1B). Similarly, LC-MS/MS identified 561 phosphoproteins in the circulating EVs, which included 421 (75%) of ExoCarta proteins (Figure 1B). These results indicate that the isolated proteome and phosphoproteome are enriched for small EVs (exosomes).

**Figure 1.**
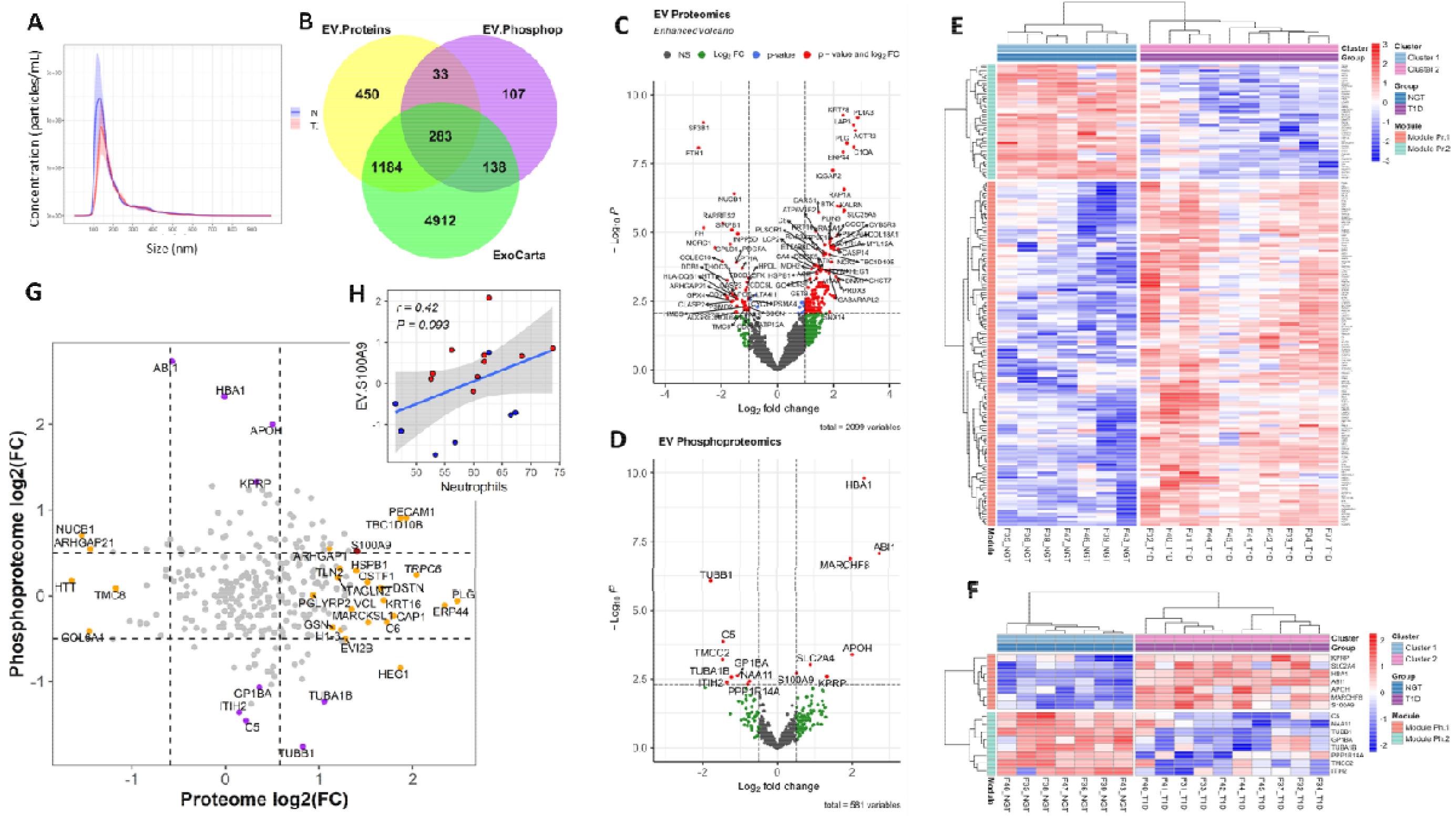
EV Characterization. A)Nanoparticle Tracking Analysis (NTA) of EVs isolated from participants with T1D and NGT. B) Overlap between the exosomal proteins reported in Exocarta and all the EV proteins and phosphoproteins identified in the present work. C,D) Volcano plots of the EV proteome and phosphoproteome. Dashed lines indicate p value = 0.0085 (nominal p that produced FDR < 0.1) and two-fold expression change (for proteomic analysis) and p value = 0.005 (nominal for FDR < 0.2) and 1.41-fold change for phosphoproteomic analysis. E-F) Unsupervised clustering of the differentially expressed proteome and phosphoproteome, respectively. G) Correlation plot of EV proteome versus phosphoproteome (grey circles: non-significant change, orange circles: differentially expressed only in proteomic analysis, purple circles: differentially expressed only in phosphoproteomic analysis; dark red circle: differentially expressed in both proteomic and phosphoproteomic analyses). H) Correlation between the S100A9 protein in circulating EVs and the number of circulating neutrophils.

### The serum EV proteome and phosphoproteome are different in people with T1D compared to controls

To further characterize the circulating EV cargo in our study population, we conducted additional bioinformatic analyses including important variable analysis with random forests (RF), differential expression analysis, and unsupervised clustering analysis. A total of 181 differentially expressed EV proteins were identified with, at least, a 2-fold change (p-value ≤ 0.01, FDR<0.1). Of these, 135 were upregulated and 46 were downregulated in T1D, as compared to controls (Table 1, Figure 1E). On the other hand, 15 EV phosphoproteins were more than 1.4-fold different between the groups (p-value < 0.05, FDR < 0.2), with 7 of them being upregulated and 8 being downregulated (Table 2, Figure 1F). Notably, of the top 100 exosomal proteins reported by ExoCarta, which are considered the best markers of exosomes, 8 (i.e., CD63, RAB14, VCP, BSG (a.k.a. CD147), FLNA, GNAI2, LAMP2, EZR), were significantly upregulated (absolute fold change FC ≥ 2, P < 0.01, FDR < 0.1), while 1 phosphoprotein (i.e., TUBA1B) was downregulated (FC = -1.75, P = 0.0027, FDR = 0.13) in the circulating EVs from the T1D group. Interestingly, the EV proteome and phosphoproteome showed a relatively small overlap, in general. Only a single protein (S100A9) was differentially expressed (i.e., upregulated) at both the total protein and the phosphorylated state (Figure 2G). All other differentially expressed proteins/phosphoproteins were only differentially expressed at either the total protein or the phosphorylated protein level. S100A9 displayed a positive trend (r = 0.42, P = 0.093) with the number of circulating neutrophils (Figure 1H).

**Table 1.**
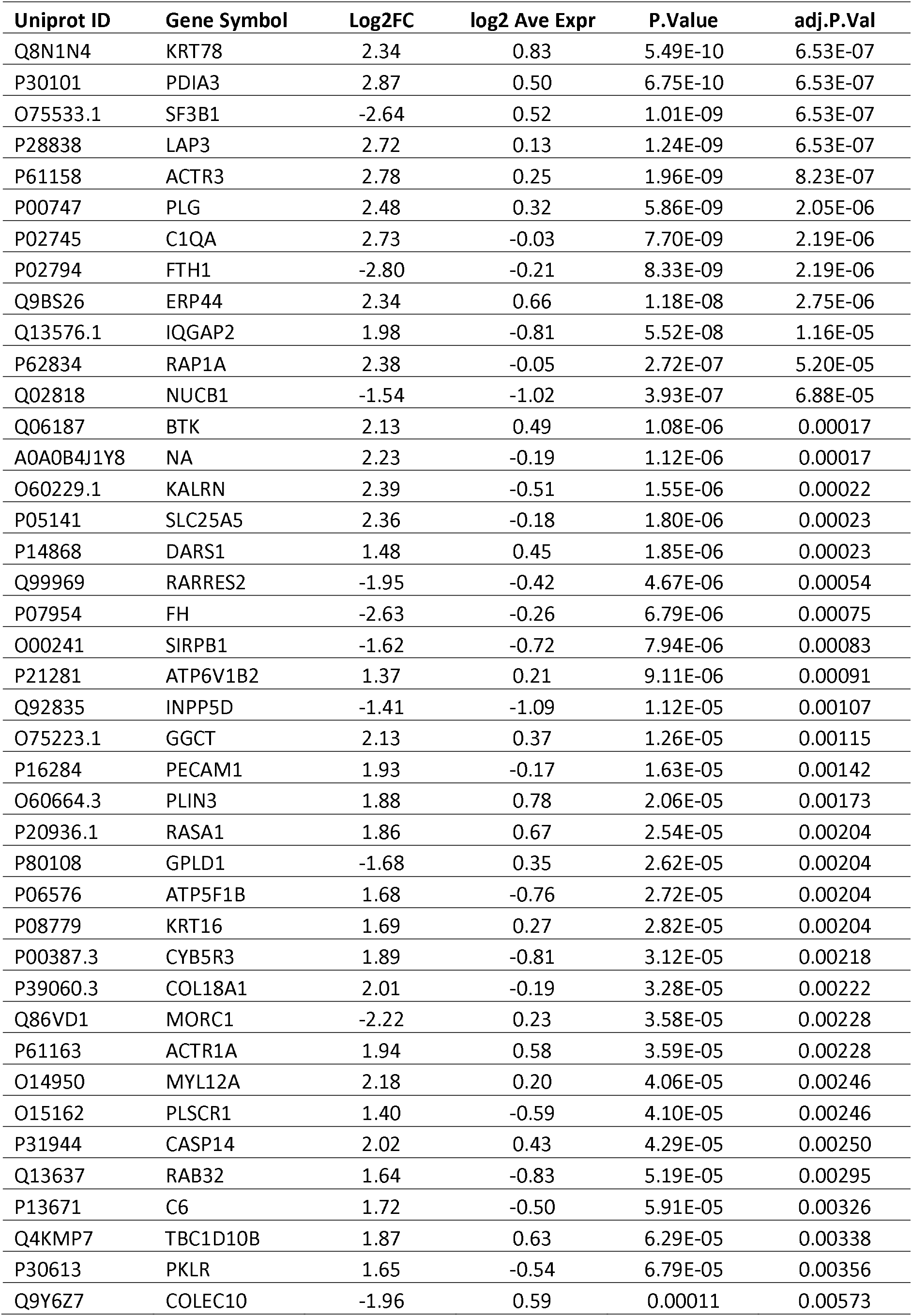
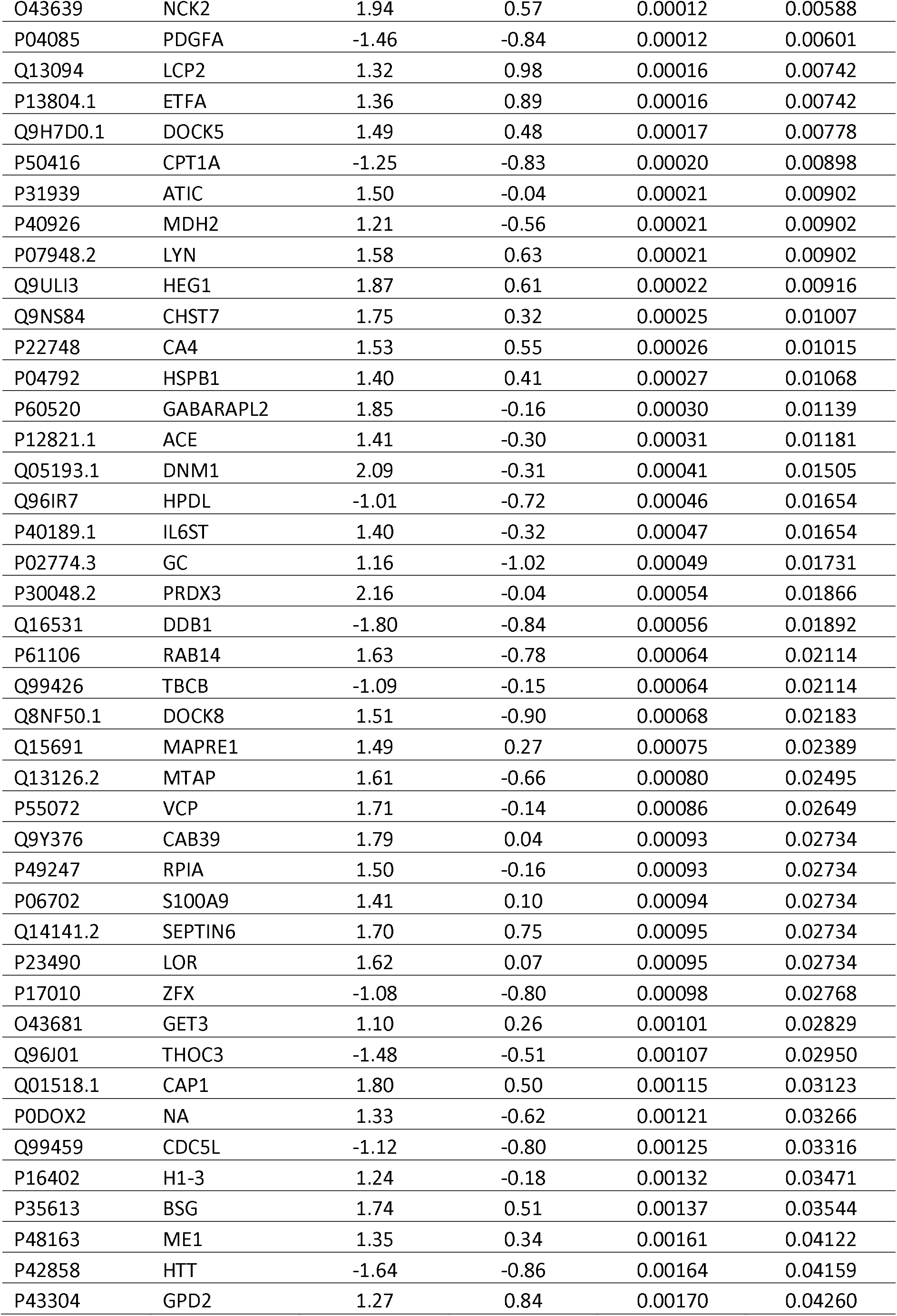
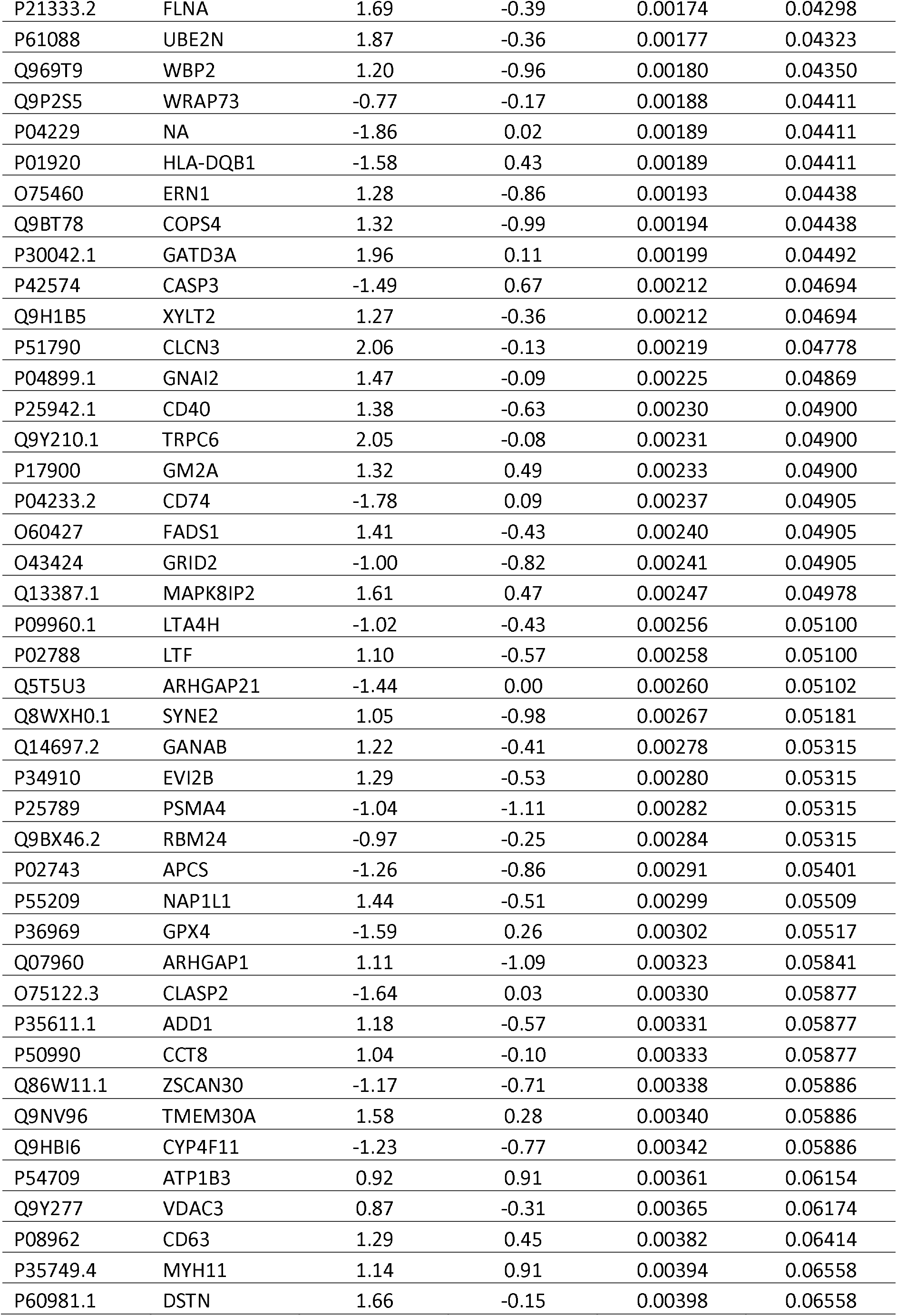
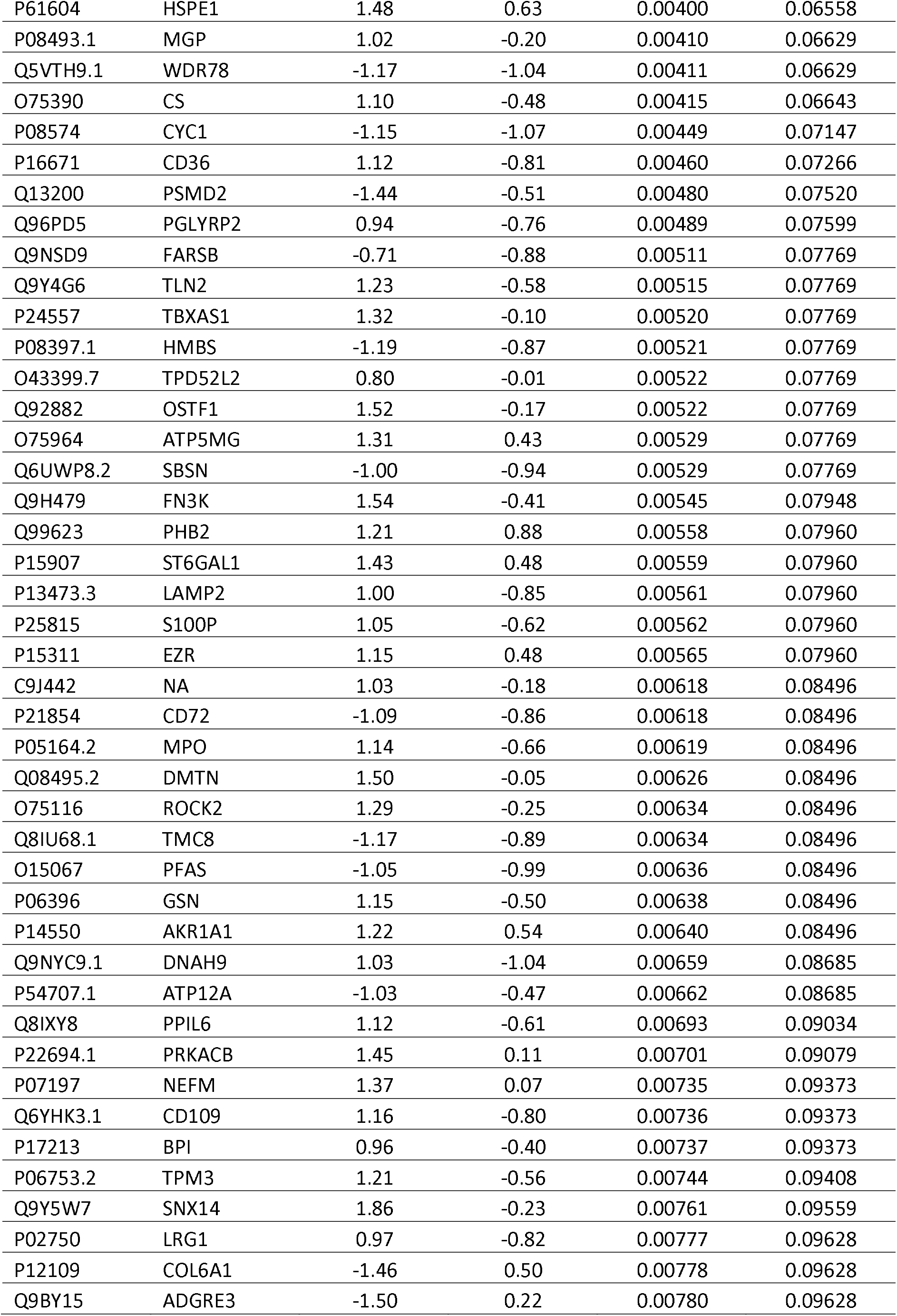
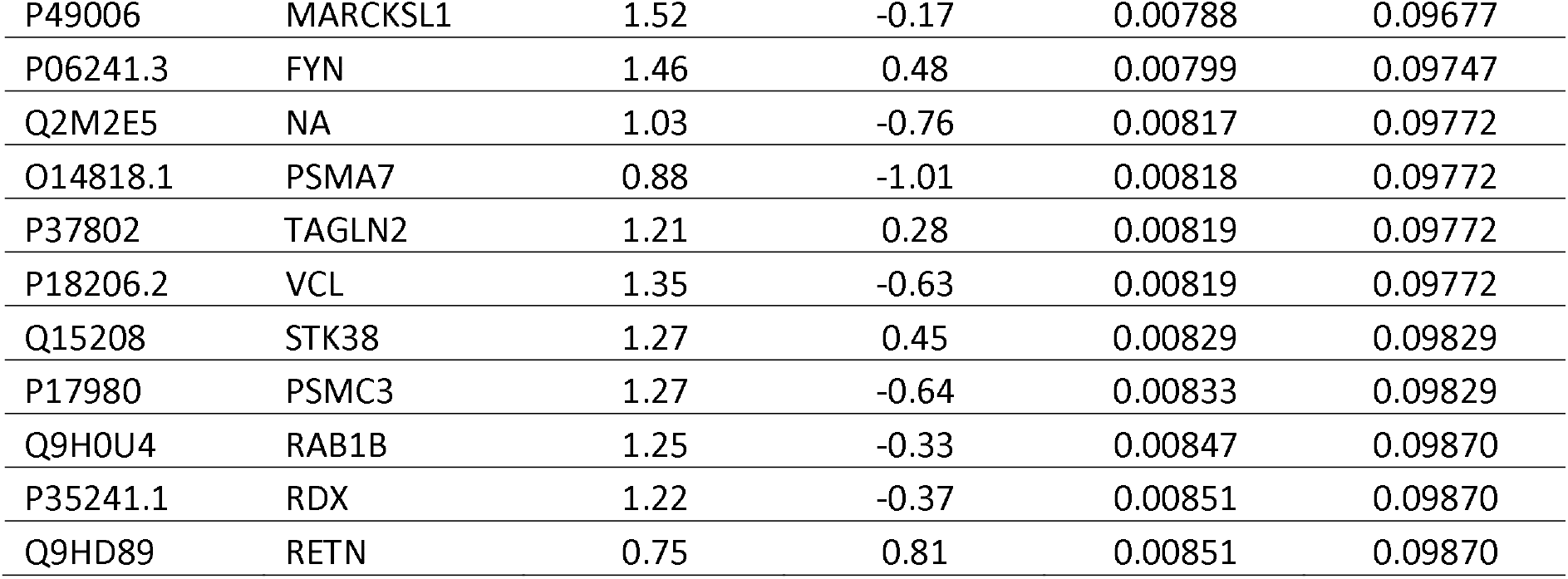
Differentially expressed proteins in circulating EVs (T1D vs. NGT)

| Uniprot ID | Gene Symbol | Log2FC | log2 Ave Expr | P.Value | adj.P.Val |
| --- | --- | --- | --- | --- | --- |
| Q8N1N4 | KRT78 | 2.34 | 0.83 | 5.49E-10 | 6.53E-07 |
| P30101 | PDIA3 | 2.87 | 0.50 | 6.75E-10 | 6.53E-07 |
| O75533.1 | SF3B1 | -2.64 | 0.52 | 1.01E-09 | 6.53E-07 |
| P28838 | LAP3 | 2.72 | 0.13 | 1.24E-09 | 6.53E-07 |
| P61158 | ACTR3 | 2.78 | 0.25 | 1.96E-09 | 8.23E-07 |
| P00747 | PLG | 2.48 | 0.32 | 5.86E-09 | 2.05E-06 |
| P02745 | C1QA | 2.73 | -0.03 | 7.70E-09 | 2.19E-06 |
| P02794 | FTH1 | -2.80 | -0.21 | 8.33E-09 | 2.19E-06 |
| Q9BS26 | ERP44 | 2.34 | 0.66 | 1.18E-08 | 2.75E-06 |
| Q13576.1 | IQGAP2 | 1.98 | -0.81 | 5.52E-08 | 1.16E-05 |
| P62834 | RAP1A | 2.38 | -0.05 | 2.72E-07 | 5.20E-05 |
| Q02818 | NUCB1 | -1.54 | -1.02 | 3.93E-07 | 6.88E-05 |
| Q06187 | BTK | 2.13 | 0.49 | 1.08E-06 | 0.00017 |
| A0A0B4J1Y8 | NA | 2.23 | -0.19 | 1.12E-06 | 0.00017 |
| O60229.1 | KALRN | 2.39 | -0.51 | 1.55E-06 | 0.00022 |
| P05141 | SLC25A5 | 2.36 | -0.18 | 1.80E-06 | 0.00023 |
| P14868 | DARS1 | 1.48 | 0.45 | 1.85E-06 | 0.00023 |
| Q99969 | RARRES2 | -1.95 | -0.42 | 4.67E-06 | 0.00054 |
| P07954 | FH | -2.63 | -0.26 | 6.79E-06 | 0.00075 |
| O00241 | SIRPB1 | -1.62 | -0.72 | 7.94E-06 | 0.00083 |
| P21281 | ATP6V1B2 | 1.37 | 0.21 | 9.11E-06 | 0.00091 |
| Q92835 | INPP5D | -1.41 | -1.09 | 1.12E-05 | 0.00107 |
| O75223.1 | GGCT | 2.13 | 0.37 | 1.26E-05 | 0.00115 |
| P16284 | PECAM1 | 1.93 | -0.17 | 1.63E-05 | 0.00142 |
| O60664.3 | PLIN3 | 1.88 | 0.78 | 2.06E-05 | 0.00173 |
| P20936.1 | RASA1 | 1.86 | 0.67 | 2.54E-05 | 0.00204 |
| P80108 | GPLD1 | -1.68 | 0.35 | 2.62E-05 | 0.00204 |
| P06576 | ATP5F1B | 1.68 | -0.76 | 2.72E-05 | 0.00204 |
| P08779 | KRT16 | 1.69 | 0.27 | 2.82E-05 | 0.00204 |
| P00387.3 | CYB5R3 | 1.89 | -0.81 | 3.12E-05 | 0.00218 |
| P39060.3 | COL18A1 | 2.01 | -0.19 | 3.28E-05 | 0.00222 |
| Q86VD1 | MORC1 | -2.22 | 0.23 | 3.58E-05 | 0.00228 |
| P61163 | ACTR1A | 1.94 | 0.58 | 3.59E-05 | 0.00228 |
| O14950 | MYL12A | 2.18 | 0.20 | 4.06E-05 | 0.00246 |
| O15162 | PLSCR1 | 1.40 | -0.59 | 4.10E-05 | 0.00246 |
| P31944 | CASP14 | 2.02 | 0.43 | 4.29E-05 | 0.00250 |
| Q13637 | RAB32 | 1.64 | -0.83 | 5.19E-05 | 0.00295 |
| P13671 | C6 | 1.72 | -0.50 | 5.91E-05 | 0.00326 |
| Q4KMP7 | TBC1D10B | 1.87 | 0.63 | 6.29E-05 | 0.00338 |
| P30613 | PKLR | 1.65 | -0.54 | 6.79E-05 | 0.00356 |
| Q9Y6Z7 | COLEC10 | -1.96 | 0.59 | 0.00011 | 0.00573 |
| O43639 | NCK2 | 1.94 | 0.57 | 0.00012 | 0.00588 |
| P04085 | PDGFA | -1.46 | -0.84 | 0.00012 | 0.00601 |
| Q13094 | LCP2 | 1.32 | 0.98 | 0.00016 | 0.00742 |
| P13804.1 | ETFA | 1.36 | 0.89 | 0.00016 | 0.00742 |
| Q9H7D0.1 | DOCK5 | 1.49 | 0.48 | 0.00017 | 0.00778 |
| P50416 | CPT1A | -1.25 | -0.83 | 0.00020 | 0.00898 |
| P31939 | ATIC | 1.50 | -0.04 | 0.00021 | 0.00902 |
| P40926 | MDH2 | 1.21 | -0.56 | 0.00021 | 0.00902 |
| P07948.2 | LYN | 1.58 | 0.63 | 0.00021 | 0.00902 |
| Q9ULI3 | HEG1 | 1.87 | 0.61 | 0.00022 | 0.00916 |
| Q9NS84 | CHST7 | 1.75 | 0.32 | 0.00025 | 0.01007 |
| P22748 | CA4 | 1.53 | 0.55 | 0.00026 | 0.01015 |
| P04792 | HSPB1 | 1.40 | 0.41 | 0.00027 | 0.01068 |
| P60520 | GABARAPL2 | 1.85 | -0.16 | 0.00030 | 0.01139 |
| P12821.1 | ACE | 1.41 | -0.30 | 0.00031 | 0.01181 |
| Q05193.1 | DNM1 | 2.09 | -0.31 | 0.00041 | 0.01505 |
| Q96IR7 | HPDL | -1.01 | -0.72 | 0.00046 | 0.01654 |
| P40189.1 | IL6ST | 1.40 | -0.32 | 0.00047 | 0.01654 |
| P02774.3 | GC | 1.16 | -1.02 | 0.00049 | 0.01731 |
| P30048.2 | PRDX3 | 2.16 | -0.04 | 0.00054 | 0.01866 |
| Q16531 | DDB1 | -1.80 | -0.84 | 0.00056 | 0.01892 |
| P61106 | RAB14 | 1.63 | -0.78 | 0.00064 | 0.02114 |
| Q99426 | TBCB | -1.09 | -0.15 | 0.00064 | 0.02114 |
| Q8NF50.1 | DOCK8 | 1.51 | -0.90 | 0.00068 | 0.02183 |
| Q15691 | MAPRE1 | 1.49 | 0.27 | 0.00075 | 0.02389 |
| Q13126.2 | MTAP | 1.61 | -0.66 | 0.00080 | 0.02495 |
| P55072 | VCP | 1.71 | -0.14 | 0.00086 | 0.02649 |
| Q9Y376 | CAB39 | 1.79 | 0.04 | 0.00093 | 0.02734 |
| P49247 | RPIA | 1.50 | -0.16 | 0.00093 | 0.02734 |
| P06702 | S100A9 | 1.41 | 0.10 | 0.00094 | 0.02734 |
| Q14141.2 | SEPTIN6 | 1.70 | 0.75 | 0.00095 | 0.02734 |
| P23490 | LOR | 1.62 | 0.07 | 0.00095 | 0.02734 |
| P17010 | ZFX | -1.08 | -0.80 | 0.00098 | 0.02768 |
| O43681 | GET3 | 1.10 | 0.26 | 0.00101 | 0.02829 |
| Q96J01 | THOC3 | -1.48 | -0.51 | 0.00107 | 0.02950 |
| Q01518.1 | CAP1 | 1.80 | 0.50 | 0.00115 | 0.03123 |
| P0DOX2 | NA | 1.33 | -0.62 | 0.00121 | 0.03266 |
| Q99459 | CDC5L | -1.12 | -0.80 | 0.00125 | 0.03316 |
| P16402 | H1-3 | 1.24 | -0.18 | 0.00132 | 0.03471 |
| P35613 | BSG | 1.74 | 0.51 | 0.00137 | 0.03544 |
| P48163 | ME1 | 1.35 | 0.34 | 0.00161 | 0.04122 |
| P42858 | HTT | -1.64 | -0.86 | 0.00164 | 0.04159 |
| P43304 | GPD2 | 1.27 | 0.84 | 0.00170 | 0.04260 |
| P21333.2 | FLNA | 1.69 | -0.39 | 0.00174 | 0.04298 |
| P61088 | UBE2N | 1.87 | -0.36 | 0.00177 | 0.04323 |
| Q969T9 | WBP2 | 1.20 | -0.96 | 0.00180 | 0.04350 |
| Q9P2S5 | WRAP73 | -0.77 | -0.17 | 0.00188 | 0.04411 |
| P04229 | NA | -1.86 | 0.02 | 0.00189 | 0.04411 |
| P01920 | HLA-DQB1 | -1.58 | 0.43 | 0.00189 | 0.04411 |
| O75460 | ERN1 | 1.28 | -0.86 | 0.00193 | 0.04438 |
| Q9BT78 | COPS4 | 1.32 | -0.99 | 0.00194 | 0.04438 |
| P30042.1 | GATD3A | 1.96 | 0.11 | 0.00199 | 0.04492 |
| P42574 | CASP3 | -1.49 | 0.67 | 0.00212 | 0.04694 |
| Q9H1B5 | XYLT2 | 1.27 | -0.36 | 0.00212 | 0.04694 |
| P51790 | CLCN3 | 2.06 | -0.13 | 0.00219 | 0.04778 |
| P04899.1 | GNAI2 | 1.47 | -0.09 | 0.00225 | 0.04869 |
| P25942.1 | CD40 | 1.38 | -0.63 | 0.00230 | 0.04900 |
| Q9Y210.1 | TRPC6 | 2.05 | -0.08 | 0.00231 | 0.04900 |
| P17900 | GM2A | 1.32 | 0.49 | 0.00233 | 0.04900 |
| P04233.2 | CD74 | -1.78 | 0.09 | 0.00237 | 0.04905 |
| O60427 | FADS1 | 1.41 | -0.43 | 0.00240 | 0.04905 |
| O43424 | GRID2 | -1.00 | -0.82 | 0.00241 | 0.04905 |
| Q13387.1 | MAPK8IP2 | 1.61 | 0.47 | 0.00247 | 0.04978 |
| P09960.1 | LTA4H | -1.02 | -0.43 | 0.00256 | 0.05100 |
| P02788 | LTF | 1.10 | -0.57 | 0.00258 | 0.05100 |
| Q5T5U3 | ARHGAP21 | -1.44 | 0.00 | 0.00260 | 0.05102 |
| Q8WXH0.1 | SYNE2 | 1.05 | -0.98 | 0.00267 | 0.05181 |
| Q14697.2 | GANAB | 1.22 | -0.41 | 0.00278 | 0.05315 |
| P34910 | EVI2B | 1.29 | -0.53 | 0.00280 | 0.05315 |
| P25789 | PSMA4 | -1.04 | -1.11 | 0.00282 | 0.05315 |
| Q9BX46.2 | RBM24 | -0.97 | -0.25 | 0.00284 | 0.05315 |
| P02743 | APCS | -1.26 | -0.86 | 0.00291 | 0.05401 |
| P55209 | NAP1L1 | 1.44 | -0.51 | 0.00299 | 0.05509 |
| P36969 | GPX4 | -1.59 | 0.26 | 0.00302 | 0.05517 |
| Q07960 | ARHGAP1 | 1.11 | -1.09 | 0.00323 | 0.05841 |
| O75122.3 | CLASP2 | -1.64 | 0.03 | 0.00330 | 0.05877 |
| P35611.1 | ADD1 | 1.18 | -0.57 | 0.00331 | 0.05877 |
| P50990 | CCT8 | 1.04 | -0.10 | 0.00333 | 0.05877 |
| Q86W11.1 | ZSCAN30 | -1.17 | -0.71 | 0.00338 | 0.05886 |
| Q9NV96 | TMEM30A | 1.58 | 0.28 | 0.00340 | 0.05886 |
| Q9HBI6 | CYP4F11 | -1.23 | -0.77 | 0.00342 | 0.05886 |
| P54709 | ATP1B3 | 0.92 | 0.91 | 0.00361 | 0.06154 |
| Q9Y277 | VDAC3 | 0.87 | -0.31 | 0.00365 | 0.06174 |
| P08962 | CD63 | 1.29 | 0.45 | 0.00382 | 0.06414 |
| P35749.4 | MYH11 | 1.14 | 0.91 | 0.00394 | 0.06558 |
| P60981.1 | DSTN | 1.66 | -0.15 | 0.00398 | 0.06558 |
| P61604 | HSPE1 | 1.48 | 0.63 | 0.00400 | 0.06558 |
| P08493.1 | MGP | 1.02 | -0.20 | 0.00410 | 0.06629 |
| Q5VTH9.1 | WDR78 | -1.17 | -1.04 | 0.00411 | 0.06629 |
| O75390 | CS | 1.10 | -0.48 | 0.00415 | 0.06643 |
| P08574 | CYC1 | -1.15 | -1.07 | 0.00449 | 0.07147 |
| P16671 | CD36 | 1.12 | -0.81 | 0.00460 | 0.07266 |
| Q13200 | PSMD2 | -1.44 | -0.51 | 0.00480 | 0.07520 |
| Q96PD5 | PGLYRP2 | 0.94 | -0.76 | 0.00489 | 0.07599 |
| Q9NSD9 | FARSB | -0.71 | -0.88 | 0.00511 | 0.07769 |
| Q9Y4G6 | TLN2 | 1.23 | -0.58 | 0.00515 | 0.07769 |
| P24557 | TBXAS1 | 1.32 | -0.10 | 0.00520 | 0.07769 |
| P08397.1 | HMBS | -1.19 | -0.87 | 0.00521 | 0.07769 |
| O43399.7 | TPD52L2 | 0.80 | -0.01 | 0.00522 | 0.07769 |
| Q92882 | OSTF1 | 1.52 | -0.17 | 0.00522 | 0.07769 |
| O75964 | ATP5MG | 1.31 | 0.43 | 0.00529 | 0.07769 |
| Q6UWP8.2 | SBSN | -1.00 | -0.94 | 0.00529 | 0.07769 |
| Q9H479 | FN3K | 1.54 | -0.41 | 0.00545 | 0.07948 |
| Q99623 | PHB2 | 1.21 | 0.88 | 0.00558 | 0.07960 |
| P15907 | ST6GAL1 | 1.43 | 0.48 | 0.00559 | 0.07960 |
| P13473.3 | LAMP2 | 1.00 | -0.85 | 0.00561 | 0.07960 |
| P25815 | S100P | 1.05 | -0.62 | 0.00562 | 0.07960 |
| P15311 | EZR | 1.15 | 0.48 | 0.00565 | 0.07960 |
| C9J442 | NA | 1.03 | -0.18 | 0.00618 | 0.08496 |
| P21854 | CD72 | -1.09 | -0.86 | 0.00618 | 0.08496 |
| P05164.2 | MPO | 1.14 | -0.66 | 0.00619 | 0.08496 |
| Q08495.2 | DMTN | 1.50 | -0.05 | 0.00626 | 0.08496 |
| O75116 | ROCK2 | 1.29 | -0.25 | 0.00634 | 0.08496 |
| Q8IU68.1 | TMC8 | -1.17 | -0.89 | 0.00634 | 0.08496 |
| O15067 | PFAS | -1.05 | -0.99 | 0.00636 | 0.08496 |
| P06396 | GSN | 1.15 | -0.50 | 0.00638 | 0.08496 |
| P14550 | AKR1A1 | 1.22 | 0.54 | 0.00640 | 0.08496 |
| Q9NYC9.1 | DNAH9 | 1.03 | -1.04 | 0.00659 | 0.08685 |
| P54707.1 | ATP12A | -1.03 | -0.47 | 0.00662 | 0.08685 |
| Q8IXY8 | PPIL6 | 1.12 | -0.61 | 0.00693 | 0.09034 |
| P22694.1 | PRKACB | 1.45 | 0.11 | 0.00701 | 0.09079 |
| P07197 | NEFM | 1.37 | 0.07 | 0.00735 | 0.09373 |
| Q6YHK3.1 | CD109 | 1.16 | -0.80 | 0.00736 | 0.09373 |
| P17213 | BPI | 0.96 | -0.40 | 0.00737 | 0.09373 |
| P06753.2 | TPM3 | 1.21 | -0.56 | 0.00744 | 0.09408 |
| Q9Y5W7 | SNX14 | 1.86 | -0.23 | 0.00761 | 0.09559 |
| P02750 | LRG1 | 0.97 | -0.82 | 0.00777 | 0.09628 |
| P12109 | COL6A1 | -1.46 | 0.50 | 0.00778 | 0.09628 |
| Q9BY15 | ADGRE3 | -1.50 | 0.22 | 0.00780 | 0.09628 |
| P49006 | MARCKSL1 | 1.52 | -0.17 | 0.00788 | 0.09677 |
| P06241.3 | FYN | 1.46 | 0.48 | 0.00799 | 0.09747 |
| Q2M2E5 | NA | 1.03 | -0.76 | 0.00817 | 0.09772 |
| O14818.1 | PSMA7 | 0.88 | -1.01 | 0.00818 | 0.09772 |
| P37802 | TAGLN2 | 1.21 | 0.28 | 0.00819 | 0.09772 |
| P18206.2 | VCL | 1.35 | -0.63 | 0.00819 | 0.09772 |
| Q15208 | STK38 | 1.27 | 0.45 | 0.00829 | 0.09829 |
| P17980 | PSMC3 | 1.27 | -0.64 | 0.00833 | 0.09829 |
| Q9H0U4 | RAB1B | 1.25 | -0.33 | 0.00847 | 0.09870 |
| P35241.1 | RDX | 1.22 | -0.37 | 0.00851 | 0.09870 |
| Q9HD89 | RETN | 0.75 | 0.81 | 0.00851 | 0.09870 |

**Table 2:** Differentially expressed phosphoproteins in circulating EVs (T1D vs. NGT)

| Uniprot ID | Log2FC | log2 Ave Expr | P.Value | adj.P.Val |
| --- | --- | --- | --- | --- |
| P69905 | 2.32 | 0.57 | 1.56E-10 | 9.09E-08 |
| Q8IZP0.8 | 2.73 | 0.26 | 8.63E-08 | 2.51E-05 |
| Q5T0T0 | 1.95 | -0.52 | 1.31E-07 | 2.53E-05 |
| Q9H4B7 | -1.76 | 0.06 | 8.95E-07 | 0.00013 |
| P01031 | -1.45 | -0.69 | 0.00013 | 0.01561 |
| P02749 | 2.00 | 0.20 | 0.00041 | 0.03994 |
| O75069.2 | -1.47 | -0.76 | 0.00061 | 0.05048 |
| P14672 | 0.88 | -0.95 | 0.00095 | 0.06887 |
| P06702 | 0.52 | -1.15 | 0.00192 | 0.12377 |
| P07359 | -1.06 | -1.04 | 0.00236 | 0.13089 |
| Q5T749 | 1.32 | -0.94 | 0.00259 | 0.13089 |
| P68363 | -1.23 | -0.63 | 0.00270 | 0.13089 |
| Q9BSU3 | -0.74 | -1.07 | 0.00380 | 0.16982 |
| P19823 | -1.36 | 0.25 | 0.00421 | 0.17476 |
| Q96A00 | -0.80 | -1.14 | 0.00479 | 0.18572 |

**Figure 2.**
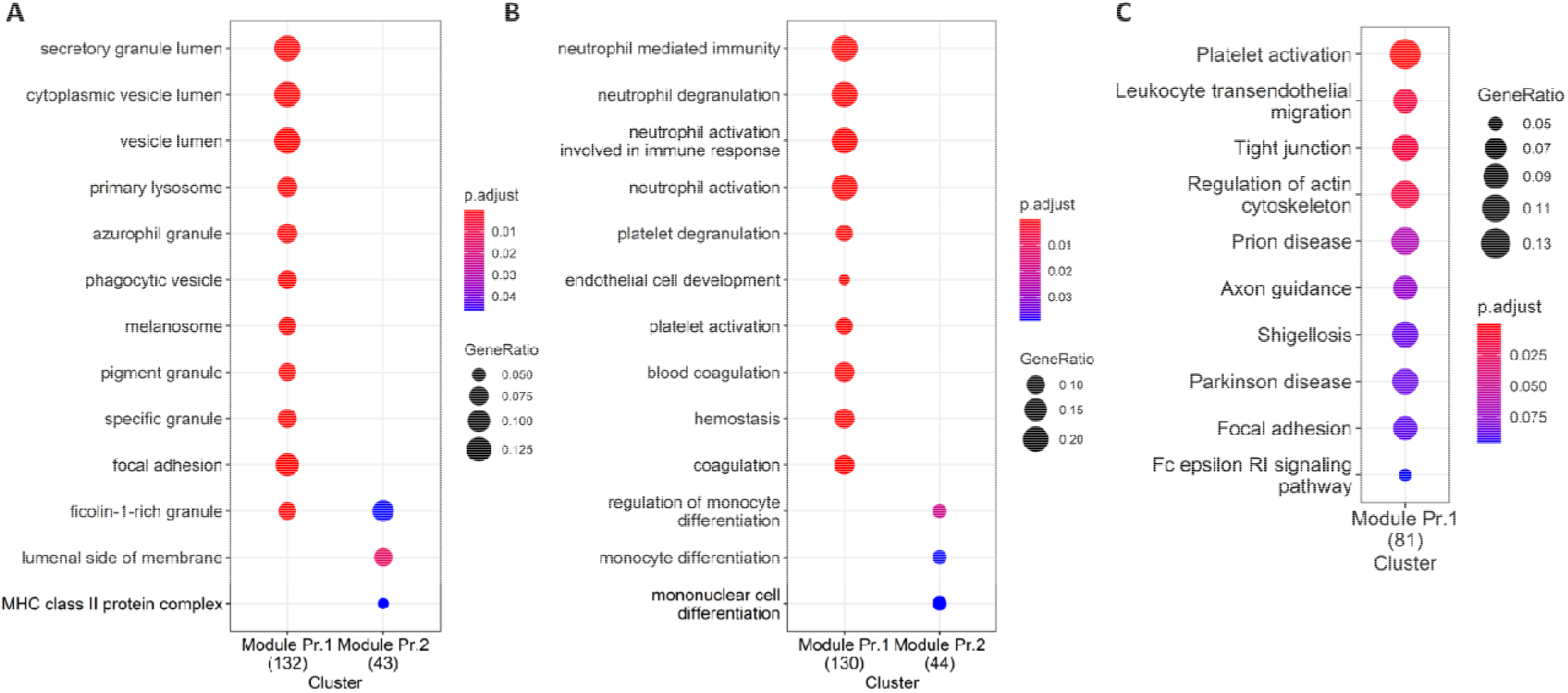
Enrichment analyses for differentially expressed EV proteins. A) GO Cellular Compartment. B) GO Biological Processes. C) KEGG pathways.

As pathway analysis techniques can help in interpreting proteomics results, we applied various enrichment analysis tools to obtain functional information on the differentially expressed proteins. The Gene Ontology (GO) cellular compartment enrichment analysis confirmed that the differentially expressed proteins primarily belong to pathways associated with vesicles and granules: secretory granule lumen, cytoplasmic vesicle lumen, vesicle lumen, membrane raft, membrane microdomain, membrane region indirectly confirming the vesicular origin of the proteins (Figure 2A). GO biological process enrichment analysis on the differentially expressed EV proteins showed that the differentially upregulated proteins are involved in neutrophil degranulation and activation, immune response, platelet degranulation, blood coagulation, and hemostasis, among others (Figure 2B, Module Pr.1). On the other hand, downregulated EV proteins were enriched in proteins involved in the regulation of monocyte differentiation (Figure 2B, Module Pr.2). The Kyoto Encyclopedia of Genes and Genomes (KEGG) pathway analysis also identified platelet activation as one of the most represented pathways together with leukocyte transendothelial migration, prion disease and neurodegenerative disorders (Figures 1C).

The differentially expressed EV phosphoproteins were similarly enriched for proteins found in vesicular compartments such as late endosomes and secretory granules apparently involved in insulin response and platelet function (Figure 3A). On the other hand, enriched GO biological processes suggest the upregulated EV phosphoproteins are involved in the detoxification of cellular oxidants (Figure 3B). The KEGG pathway analysis demonstrated enrichment in upregulated EV phosphoproteins involved in insulin resistance and diabetes (Figure 3C, Module Ph.1), while downregulated EV phosphoproteins were involved in phagosome function, prion disease and neurodegenerative disease pathways (Figure 3C, Module Ph.2).

**Figure 3.**
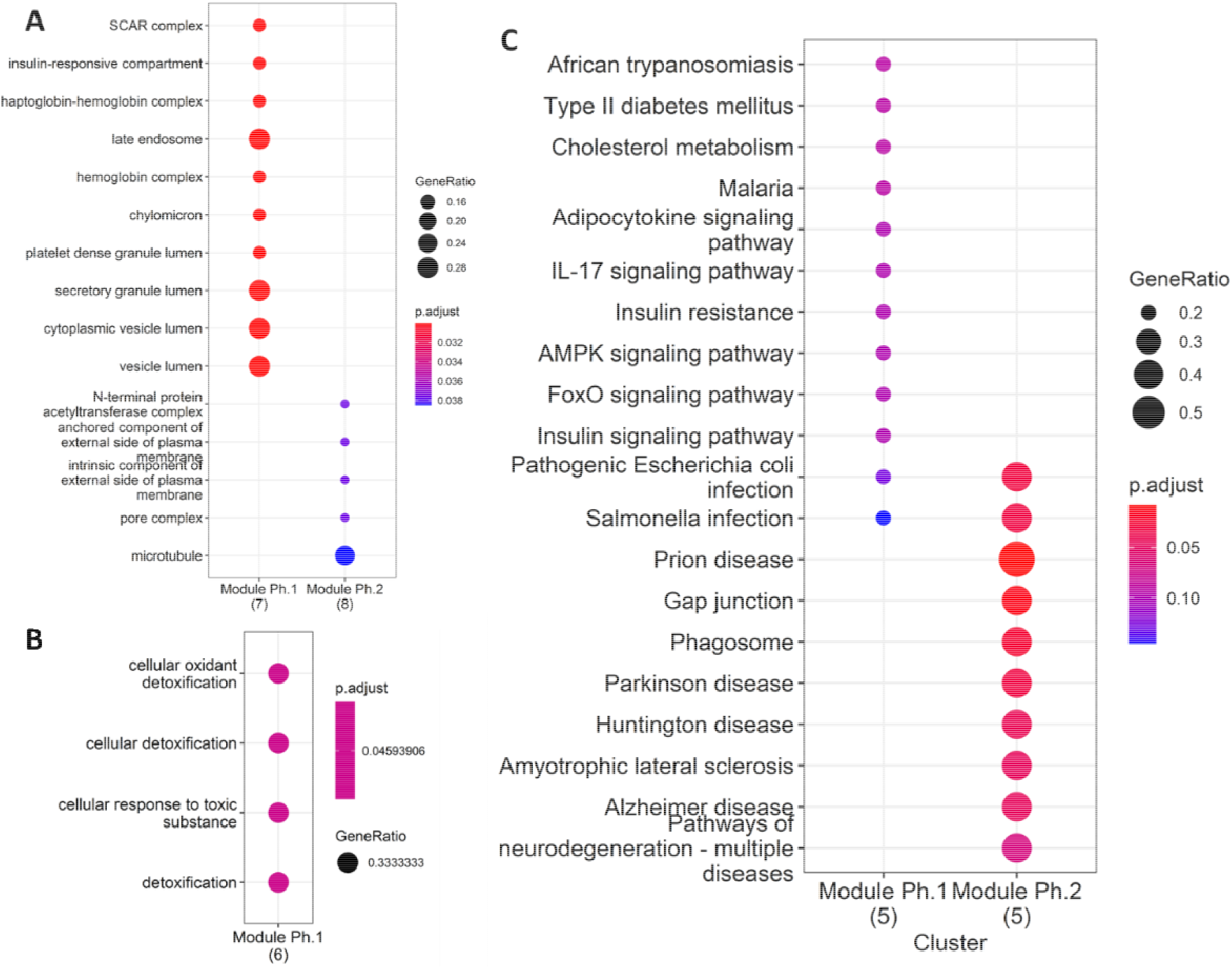
Enrichment analyses for differentially expressed EV phosphoroteins. A) GO Cellular Compartment. B) GO Biological Processes. C) KEGG pathways.

### Weighted gene co-expression network analysis identified key EV-associated protein modules that correlate with pancreas size

To further improved our understanding of the role of circulating EVs in T1D at the system level, we applied the weighted gene co-expression network analysis (WGCNA) methodology for the analysis of all proteins identified in circulating EVs. This method focuses on the identification of modules that include features (i.e., EV proteins in our case) with correlated expression patterns as opposed to the identification of individual genes based on group average comparisons (as is the case of differential expression analysis), therefore, alleviating the multiple statistical testing problem. A Pearson correlation matrix of the proteins is used to form a hierarchical dendrogram that is then cut into branches corresponding to modules. Each module includes genes with similar expression pattern and most likely specific biological functions. The module eigengenes are also correlated with external clinical traits, consequently generating a denser mechanistic overview of co-regulated EV proteins associated with the underlying T1D biology. The WGCNA methodology has been previously used for proteomic analysis (21, 22).

Figure 4A presents the co-expression dendrograms for all EV proteins and how they were grouped in 14 color-coded modules. Figure 4B presents the number of EV proteins included in each module. The analysis identified significant associations of specific EV protein modules with 9 of the 154 clinical characteristics included. Primarily, the association was with indicators of metabolic compensation and glucose levels, but other interesting traits were identified as an index of insulin sensitivity (eIS), pancreas size and resting energy expenditure. The strongest correlations were seen with clinical traits associated with blood glucose levels (metrics obtained from the continuous glucose monitoring, HbA1c) but also with other clinical characteristics indicative of T1D (pancreas size) or metabolic aspects known to be modified in T1D (Figure 4C).

**Figure 4.**
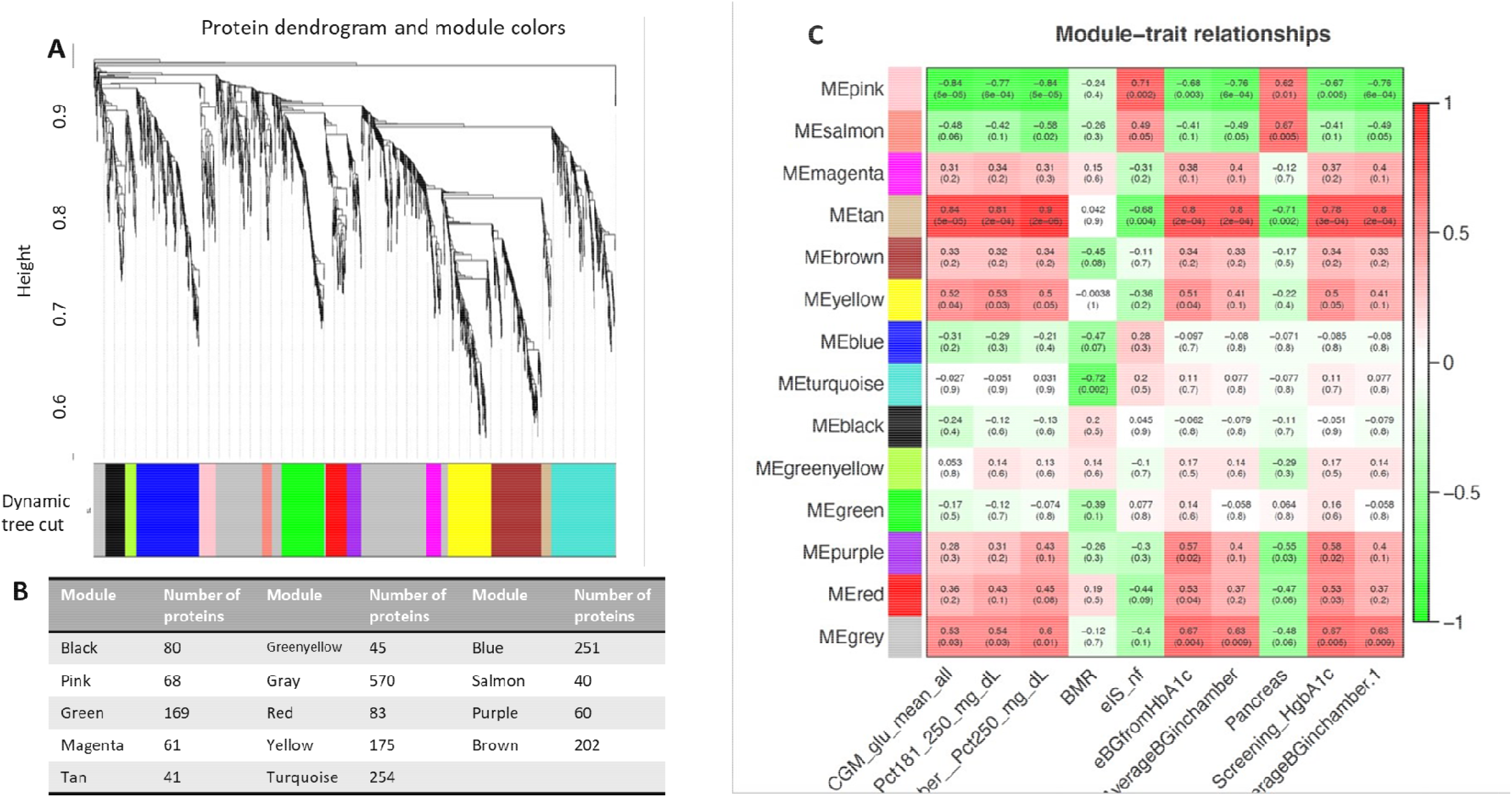
A) Protein dendrogram and module colors. B) Module size. C) Relationships among modules and clinical traits (14 modules 163 clinical trait). Row corresponds to a consensus module. Columns correspond to a trait. Numbers in the table report the correlations of the corresponding module eigengenes and traits, with the p-value below in parenthesis.

Three co-expression modules (i.e., Pink, Tan, and Yellow modules) were significantly associated with elevated blood glucose levels, expressed as either percentage of continuous glucose monitoring (CGM) above 250 mg/dl or between 180 and 250 mg/dl, average glucose level during CGM, HbA1c. As this is the characteristic feature of diabetes it is hard to discriminate whether the EV proteins included in these modules are differentially abundant as a cause or a consequence of hyperglycemia (Figure 4C). The Tan module was of particular interest because, in addition to its association with blood glucose levels (correlation 0.9 with % CGM reading >250 mg/dl, p =2e^-08^), it is associated with pancreas size (correlation -0.71, p=0.002) and estimated insulin sensitivity calculated from non-fasting parameter by the Coronary Artery Calcification in Type 1 Diabetes (CACTI) study (eIS-nf) (23).

Four modules (i.e., Tan, Salmon, Pink, Purple, Figure 4C and Figure 5) showed significant association with pancreas size that may be a pathogenic clinical feature of T1D (24-27). Heatmaps of select module proteins (36 of 41 in the Tan module and 19 of 40 in the Salmon module) that displays strong correlations between module membership and gene significance demonstrate the specificity of the EV protein-pancreas size associations with T1D (Figure 5C,D), particularly for the Tan module EV proteins. The Pink module also showed a significant correlation between module membership and gene significance. However, the association was not significant for the purple module (Figure 5A). The GO cellular component enrichment analysis for the Tan module showed a significant enrichment for membrane raft, membrane microdomain, and membrane region compartments as the highest positively correlated components. Among the enriched GO biological processes, several processes associated with inflammation and the immune response were identified (Figure 5E). The EV proteins belonging to the Salmon module and associated with pancreas were enriched for humoral immune response, regulation of viral entry into host cells, modulation by symbiont of entry into host, regulation of viral life cycle, movement in host environment, interaction with host, regulation of viral processes (Figure 5F) that might be relevant in T1D pathogenesis or response to the environment.

**Figure 5.**
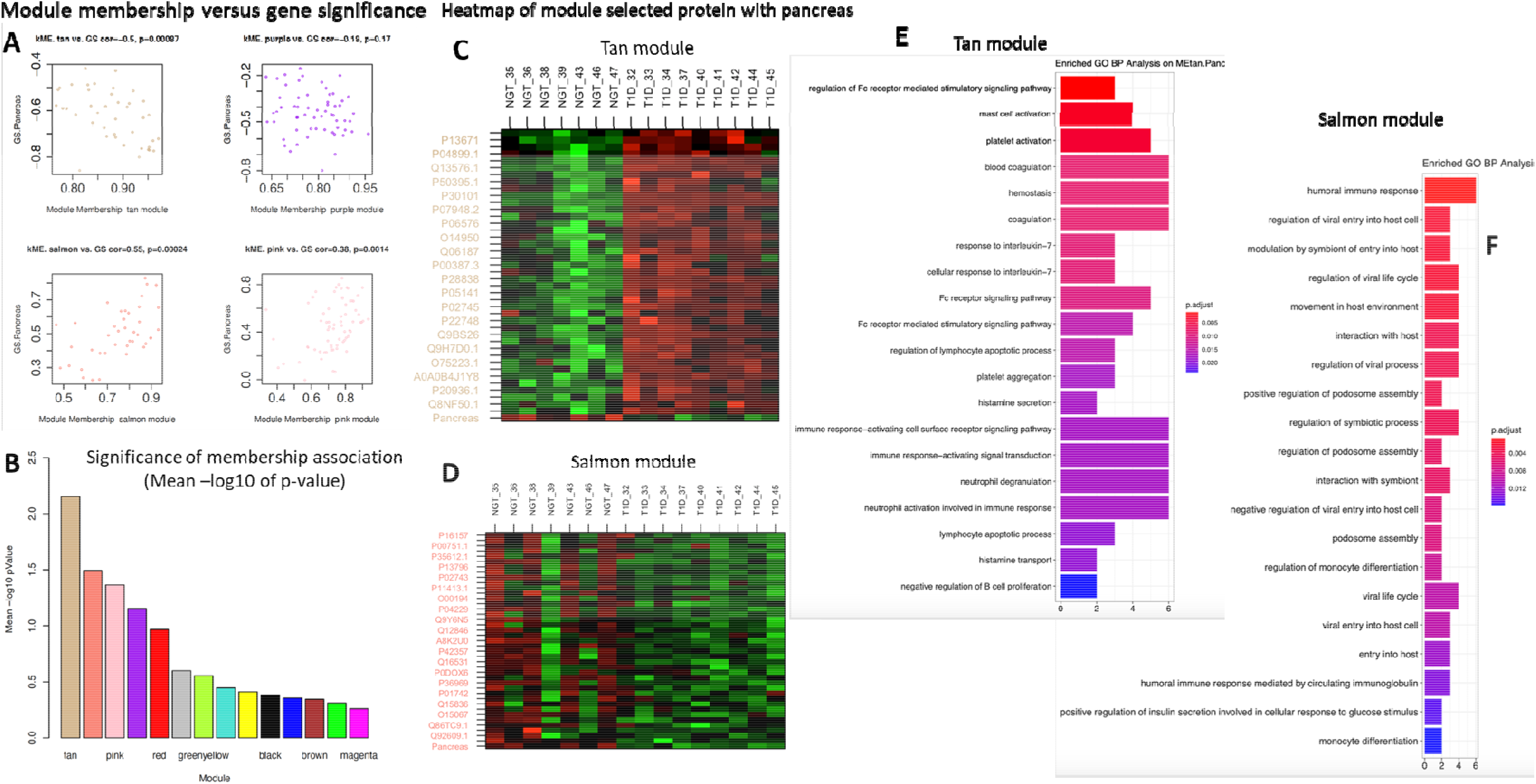
Module connectivity with pancreas size. A) Distribution of module membership versus gene significance. B) Significance of membership association with the trait of interest (pancreas size). C) Heatmap of the EV proteins correlation with pancreas size in the Tan module. D) Heatmap of the EV proteins correlation with pancreas size in the Salmon module. E and D) Enriched GO BP analysis of selected proteins associated with pancreas size in the Tan module (E) and in the Salmon module (F).

Considering the proteins with the strongest correlation with pancreas size in the two modules, we noticed that they are continuously distributed rather than separated in two groups corresponding to T1D and controls (Figure 6). This could indicate that the correlation is with the organ size rather than being mediated by a confounder that would discriminate people with diabetes such as glycemic control. The Salmon module identified proteins strongly correlated with the clinical trait of interest (i.e., pancreas size) but not appearing among the significantly differentially expressed proteins. Two of these are HLA-DRB1 and CD74 (stabilizes the MHC Class II antigen processing and prevents loading antigen peptides into the MHC Class II complex). Both HLA-DRB1 and HLA-DQB1 in serum EVs are positively associated with pancreas size and are downregulated in T1D compared to controls. This supports the concept that more sophisticated analyses are needed to understand the complex picture of circulating EV proteins.

**Figure 6.**
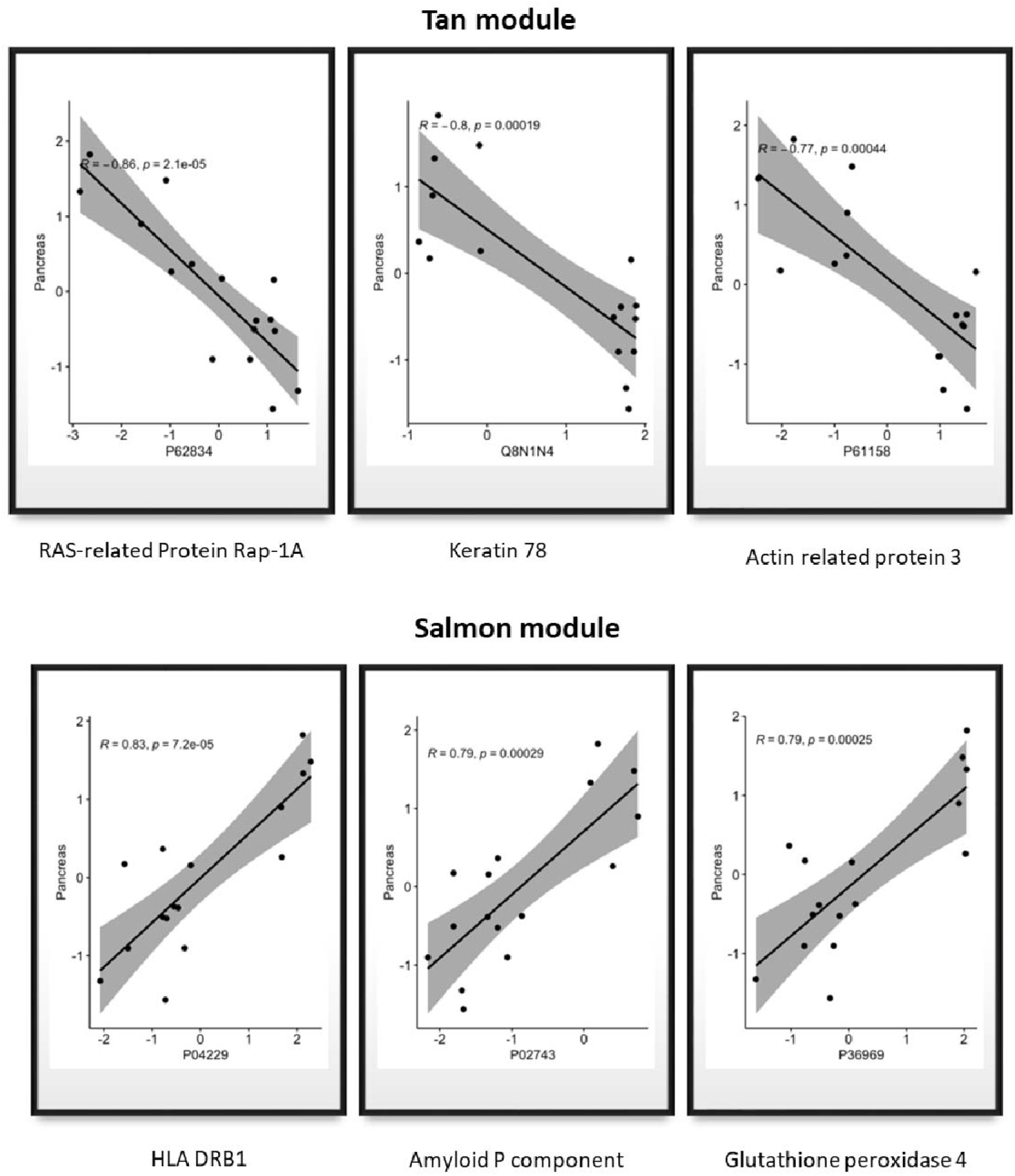
A) Proteins with a significant correlation with pancreas size in Tan module. B) Proteins with a significant correlation with pancreas size in Salmon module.

## Discussion

This pilot study showed that the circulating EV proteome significantly correlates with key features of type 1 diabetes. Rather than looking at freely circulating peripheral blood biomarkers, our study focused on the proteomic cargo in circulating EVs. The EV cargo is reported to mirror, to some extent, the specific cellular phenotypes from their cells of origins (28). Therefore, it could provide information on mechanisms that are active in the disease of interest. The importance of EV proteins as disease biomarkers has been highlighted in the cancer field (29). To our knowledge, this is the first characterization of the proteome of circulating EVs in patients with T1D compared to controls. The plasma or serum proteome includes a heterogenous pool of EVs derived by multiple cell types, with production driven by multiple mechanisms (30, 31). We would expect, therefore, that the EV proteome and phosphoproteome would be affected by hyperglycemia, insulin treatment and glucose variability as well as T1D pathogenic mechanisms. Our proteomic and phosphoproteomic analyses of serum EVs isolated using the EVtrap technology demonstrated significant enrichment in annotated exosomal proteins and phosphoproteins. This suggests that our study preparations were likely enriched in exosomes.

Supporting the information richness and biomarker potential of circulating EVs, our bioinformatic approaches were able to discriminate the two study populations, namely people with T1D and controls, and to identify key EV proteins that represent aspects of validated and novel T1D biology. By analyzing cellular compartment annotations for the differentially expressed EV proteins and phosphoproteins, we demonstrated enrichment in vesicle and secretory granule associated proteins likely involved in insulin, platelet, and neutrophil function. This leads us to the hypothesis that T1D could indeed modulate EV biogenesis and secretion. As expected, biological process and pathway analysis identified enrichment for pathways (e.g., neutrophil degranulation and activation) that may be associated not only with the pathogenesis of the disease but also with the development of diabetic chronic complications, both at the macro and microvascular level. Pathways connected to platelet function and coagulation call for the possible use of the EV proteomics in the prediction of disease complications. Specifically, C1q and plasminogen (PLG) were two of the key complement proteins strongly increased in our T1D EV preparations. Notably, deposition of complement proteins like C1q in the EVs has been reported to be needed for complement activation and, when deregulated, to contribute to pathological states (32). These particles loaded with complement regulators are suggested to contribute to the stimulation and inhibition of T-cell responses and the interaction among immune cells (33). Furthermore, C1q has been recognized as an important pattern recognition receptor that diverts autoantigen containing extracellular vesicles from immune recognition (34). PLG, on the other hand, is reported to be more efficiently activated by PLG activators when associated with cell surfaces, as compared to the reaction in solution (35). By the same token, PLG on vesicle surfaces may be more efficiently converted into plasmin, consequently promoting fibrinolysis and immune cell modulation with systemic reach.

Additionally supporting the concept that T1D could modulate EV biogenesis and secretion, 8 of the top 100 ExoCarta proteins (considered the best exosomal markers) were found to be significantly upregulated in the T1D circulating EVs. Among those proteins, CD63, a classical surface marker of exosomes, has recently being implicated as a key molecule that determines the destination of nascent insulin granules towards degradation via a novel lysosomal-mediated mechanism operative in pancreatic β cells (36). This CD63-requiring (metabolic) stress-induced nascent granule degradation (SINGD) pathway contributes to loss of insulin, β cell failure and the pathogenesis of diabetes, and its inhibition delays the onset of diabetes (36). Notably, LAMP-2, another T1D upregulated top100 ExoCarta EV protein, is also reported to interact with proteins related to glucose and lipid metabolism (37) and implicated in CD63-mediated SINGD (36). RAB14, on the other hand, has been implicated as a key regulator of GLUT4 sorting to specialized transport vesicles that translocate the glucose transporter to the plasma membrane, therefore regulating adipocyte glucose uptake (38). Filamin-A (FLNA), a protein involved in actin filament cross-linking, cell growth, and motility, has been recently implicated in insulin and IGF1 signaling (39). BSG/CD147, a membrane-bound glycoprotein involved in energy metabolism, has also been reported to be induced in monocytes by high glucose and to possibly play a role in diabetic complications (40).

Among other top upregulated EV proteins involved in the inflammatory response and immune-activation we found IL6ST/Gp130 (the founding member of the cytokine receptor family, which is involved in the regulation of adipocyte development and function and has a key role at the intersection of inflammation, autoimmunity and cancer) (41, 42) and CD40, and the HLA-DQB1 among the downregulated proteins. Of note, all these proteins are involved in IL-6 signaling (41-47). Importantly and supporting our findings, it is known that macrophages from T1D patients with high-risk HLA-DQB1 alleles are sensitized to secrete IL-6 in response to nonantigenic stimulation (43).

In addition to the neutrophil, platelet, and immune cell activation related pathways, the differentially expressed proteins and phosphoproteins were enriched for other biological processes and KEGG pathways related to prion disease and multiple neurodegenerative diseases. In this context, it is of interest to highlight that recently, the Wasserfall group (48) showed an increased frequency of cellular prion proteins in pancreas specimens from people with T1D and that, earlier, Strom and collaborators (49) demonstrated in two rodent models that altered metabolism of cellular prion protein in β cells associated with glucose dysregulation.

Only one of the differentially expressed EV proteins was also significantly differentially expressed in its phosphorylated state in circulating EVs. In this respect, S100A9, the only feature demonstrating significant differential expression (upregulation) at both the proteome and phosphoproteome level, is a determinant of the epigenetic regulation and activation of the monocyte-macrophage system in hyperglycemia (50). S100A9 in association with S100A8, form the heterocomplex known as calprotectin that is suggested to activate the transendothelial accumulation of monocytes at the site of inflammation, which associates with chronic inflammatory diseases (51, 52). S100A9 by itself is also reported to stimulates neutrophil adhesion by activating the CD11b/CD18 β2 integrin (53). Remarkably, only the phosphorylated form of S100A9 from neutrophils was shown to be essential to induce proinflammatory cytokine secretion by extracellular S100A8/phosphoS100A9 via toll-like receptor 4 signaling (54). More recently, Liao and collaborators demonstrated that S100A9 was overexpressed in pancreatic cancer-associated diabetes (PCDM) and that, together with Galectin-3, caused insulin resistance and distinguished PCDM from type 2 diabetes in subjects with new-onset diabetes (55). We now show that these processes may be additionally mediated by the presence of upregulated levels of phosphorylated S100A9 in circulating EVs.

Taking advantage of the deep metabolic phenotyping of our participants, we attempted at more precisely characterize the EV proteome and its association with clinical features (beyond the analysis of differential expression based on group averages comparisons) by applying a WGCNA methodology. Coexpression networks are powerful tools used in bioinformatics and WGCNA is a method that identifies clusters of genes based on their correlated expression patterns. The method allows for the calculation of module membership and gene significance measures relevant to the identification of biomarkers of interest associated with specific clinical variables. WGCNA has been applied in various biological contexts and “can be used to generate testable hypotheses for validation in independent data sets” (56). Thus, it aligns with the objective of our pilot study. This approach can find potential novel candidates based on co-expression similarities, rather than focusing exclusively on databases of protein-protein interaction that may have limitations due to the heterogeneity of experiments and model organisms (57). Therefore, the adaptation WGCNA on proteomic data seems particularly suitable to study EV proteomics where the literature is relatively limited and only applied to urinary EVs (58, 59).

WGCNA applied to EV proteomics in our study identified 14 distinct modules significantly associated with nine clinical features. The association is primarily with blood glucose measures indicative of T1D, but also with other features like pancreas size, resting energy expenditure, and the CACTI index of insulin sensitivity. The enrichment analysis of gene ontologies pointed out to the biological meaning of the modules, identifying neutrophil activation and degranulation, activation of immune response and blood coagulation as some of the pathways associated with hyperglycemia. Humoral immune response, regulation of viral entry and life cycle regulation of viral processes were associated with pancreas size in the Salmon module together with mast cell activation, platelet activation, blood coagulation, hemostasis, and response to IL-7 in the Tan module.

The WGCNA methodology has been applied to peripheral blood gene expression in T1D and controls (60, 61) and identified significantly disrupted co-expression modules in T1D compared to healthy control PBMCs. The module associated with T1D in Lu et al. work (60) was enriched for genes belonging to the “regulation of immune response” pathway. Our study took a broader approach considering EV proteomics. Circulating EVs originate from different cells, therefore, the modules that we identified are likely a composite of coregulated protein expressions from a number of different cellular sources, including PBMCs. In line with what was seen in PBMC transcriptomics, we also identified enrichment in proteins classified under the broad biological process ontology denominated “regulation of immune response” (that includes 1019 genes) and associated with hyperglycemia as an indicator of diabetes (regulation of Fc receptor mediated stimulatory signaling pathway). In our approach, we include proteomic profiles obtained from patients with T1D and healthy controls to identify clinically-relevant protein clusters (modules) that correlate with clinical variables of interest, independently of the disease. Therefore, the proteins included in the modules of interest could be used as markers of that clinical variable rather than of the disease. The analysis focused then on the association with pancreas size, as a trait that may be associated with disease pathogenesis.

Among the proteins of interest identified by our traditional approach (differential expression) and by WGCNA, HLA-DRB1 and DQB1 are downregulated on EVs. MHC-Class II molecules are present in exosomes originating from APC, B-cells and T-cells (62-64). Consistent with our findings, MHC Class II-loaded EVs are also detected in plasma (65). The MHC Class II in EVs is the predominant form of MHC Class II detected in serum and plasma, even though a soluble form has been described. Both the soluble MHC-Class II and the exosome-associated MHC-Class II are immunomodulatory (66). Immunosuppressive EVs derived from APCs occurs naturally, for example, soon after eating or inoculation of specific antigens (67). The experiments by Kim and collaborators (65) suggested that the immune response to a foreign antigen is regulated by exosomes in plasma produced by monocytes/macrophages that have the ability to suppress the immune response in an antigen-specific manner. Therefore, we could hypothesize that lower levels of EV MHC in T1D compared to healthy individuals could be a specific sign of the loss of tolerance in T1D. Our study is not able to demonstrate that this mechanism is active in T1D pathogenesis, however, it highlights the power of WGCNA in analyzing complex diseases and in formulating hypothesis. Paired with the in-depth phenotyping of the participants of this pilot study, WGCNA allowed us to identify and confirm the potential role of selected proteins that would have been excluded by simple differential expression analysis.

The primary limitation of our study is the small sample size. The power of the applied methodology is that it produces meaningful results even with a small sample size as in our case. The gene connectivity analysis smooths the variability and alleviates the need for multiple sampling. The design needs to be considered hypothesis generating and the findings should be confirmed in larger and independent cohort(s), and ultimately in dedicated functional studies.

## Data Availability

All data produced in the present study are available upon reasonable request to the authors

## Acknowledgements

The authors thank all ACME participants and AH/TRI staff.

## Author contributions

A.C. designed and supervised experiments, researched data, and wrote the manuscript; Y.O.N.L. conducted experiments, researched data, and wrote the manuscript; G.Y. researched data and contributed to manuscript writing; C.C. researched data and contributed to manuscript editing; A.B, contributed to clinical study execution and researched data and contributed to manuscript editing, A.M.P. conducted experiments and contributed to manuscript editing; H.C. conducted experiments and contributed to manuscript editing; K.C. conducted experiments and contributed to manuscript editing, A.I. conducted experiments, researched data, and contributed to manuscript editing; D. M. designed the larger clinical study, acquired funding, and reviewed the manuscript, E.J. M-D designed the larger clinical study, acquired funding, and reviewed the manuscript, R.E.P. designed the study, acquired funding, provided the study samples, supervised the work, and reviewed/edited the manuscript.

---

**Supplementary Table ST1:** Clinical characteristics of the study cohort

|  | **T1D (N=10)** | **NGT (N=7)** | **p** |
| --- | --- | --- | --- |
| Sex (%) |  |  | 1 |
| F | 6 (60.0) | 5 (71.4) |  |
| M | 4 (40.0) | 2 (28.6) |  |
| Race (%) |  |  | 0.245 |
| AS | 0 (0.0) | 1 (14.3) |  |
| BL | 1 (10.0) | 2 (28.6) |  |
| WH | 9 (90.0) | 4 (57.1) |  |
| Ethnic.Group (%) |  |  | 0.279 |
| H | 4 (40.0) | 4 (57.1) |  |
| NH | 6 (60.0) | 2 (28.6) |  |
| UNK | 0 (0.0) | 1 (14.3) |  |
| Age | 25.35 [24.50, 28.23] | 23.30 [23.00, 26.05] | 0.328 |
| Height_Avg | 163.50 [158.40, 168.50] | 168.60 [152.02, 171.15] | 0.696 |
| Weight_avg | 73.35 [63.15, 79.07] | 62.90 [54.95, 88.46] | 0.77 |
| BMI | 27.20 [24.12, 27.80] | 28.90 [21.80, 30.45] | 0.769 |
| SBP_avg | 126.25 [114.88, 132.00] | 120.50 [111.50, 124.25] | 0.241 |
| DBP_avg | 76.00 [71.00, 79.38] | 66.50 [62.75, 78.50] | 0.329 |
| HR_avg | 75.25 [69.00, 83.12] | 62.50 [59.00, 67.00] | 0.011 |
| Waist_avg | 86.38 [78.72, 92.25] | 78.25 [75.08, 95.50] | 0.696 |
| Hip_avg | 95.12 [91.96, 102.67] | 102.00 [85.75, 103.22] | 0.922 |
| WstHipR | 0.86 [0.84, 0.90] | 0.86 [0.81, 0.89] | 0.695 |
| Hgb.A1c | 8.10 [6.95, 9.55] | 5.10 [4.95, 5.30] | 0.001 |
| Glucose.Lvl | 142.00 [120.50, 199.50] | 90.00 [86.00, 93.50] | 0.04 |
| A.G.Ratio | 1.55 [1.33, 1.60] | 1.70 [1.40, 1.80] | 0.324 |
| Albumin.Lvl | 4.45 [4.32, 4.50] | 4.60 [4.35, 4.70] | 0.3 |
| Alk.Phos | 70.00 [62.75, 87.50] | 54.00 [49.50, 62.50] | 0.04 |
| ALT | 15.00 [11.75, 16.00] | 16.00 [13.00, 24.50] | 0.524 |
| AST | 20.00 [18.25, 22.00] | 24.00 [22.00, 26.50] | 0.171 |
| Bilirubin.Total | 0.55 [0.43, 0.75] | 0.40 [0.30, 0.45] | 0.029 |
| BUN.Lvl | 12.00 [11.00, 12.75] | 11.00 [10.00, 13.00] | 0.73 |
| Calcium.Lvl | 9.35 [9.30, 9.40] | 9.70 [9.55, 9.95] | 0.06 |
| Chloride.Lvl | 100.50 [99.25, 102.00] | 102.00 [101.00, 102.00] | 0.21 |
| Creatinine.Lvl | 0.76 [0.68, 0.83] | 0.74 [0.70, 0.85] | 0.922 |
| GFR.African.Amer.by.CKD.EPI | 139.00 [136.00, 142.00] | 135.00 [127.00, 138.75] | 0.462 |
| GFR.Non.Afr.Amer.by.CKD.EPI | 119.50 [115.25, 123.50] | 114.00 [103.00, 122.00] | 0.464 |
| Globulin.Lvl | 3.00 [2.73, 3.25] | 2.80 [2.60, 3.25] | 0.589 |
| Potassium.Lvl | 4.10 [4.00, 4.47] | 4.50 [4.30, 4.90] | 0.077 |
| Sodium.Lvl | 139.50 [138.25, 141.75] | 140.00 [138.00, 141.00] | 0.843 |
| Total.Protein.Lvl | 7.20 [7.10, 7.60] | 7.40 [7.30, 7.90] | 0.327 |
| Abs.Basophil.Cnt | 0.04 [0.04, 0.06] | 0.04 [0.03, 0.06] | 0.446 |
| Abs.Eosinophil.Cnt | 0.11 [0.09, 0.19] | 0.13 [0.07, 0.40] | 0.769 |
| Abs.Lymphocyte.Cnt | 1.54 [1.39, 1.99] | 2.01 [1.73, 2.50] | 0.13 |
| Abs.Monocyte.Cnt | 0.42 [0.36, 0.49] | 0.37 [0.34, 0.54] | 0.922 |
| Abs.Neutrophil.Cnt | 3.74 [2.84, 4.78] | 3.85 [2.80, 5.40] | 0.922 |
| Basophils | 0.70 [0.60, 0.98] | 0.50 [0.50, 0.85] | 0.183 |
| Eosinophils | 1.65 [1.35, 3.38] | 1.60 [1.25, 5.95] | 0.883 |
| Hct | 41.55 [39.42, 44.35] | 41.10 [37.65, 43.85] | 0.696 |
| Hgb | 13.80 [13.30, 14.92] | 13.00 [11.85, 14.55] | 0.328 |
| Lymphocytes | 29.10 [27.55, 30.20] | 33.60 [26.30, 38.40] | 0.435 |
| Monocytes | 7.30 [7.10, 7.68] | 5.70 [5.55, 7.40] | 0.24 |
| Neutrophils | 61.30 [57.15, 62.50] | 56.80 [50.40, 64.60] | 0.525 |
| Platelet.Count | 290.00 [254.25, 322.00] | 280.00 [248.50, 290.50] | 0.77 |
| RBC | 5.00 [4.58, 5.22] | 4.46 [4.39, 5.08] | 0.261 |
| WBC | 6.24 [4.88, 7.01] | 6.53 [5.44, 8.71] | 0.495 |
| TSH | 1.95 [1.39, 2.69] | 1.80 [1.27, 2.21] | 0.626 |
| Cholesterol | 142.50 [126.75, 167.00] | 159.00 [141.00, 168.50] | 0.329 |
| Chol.HDL.Ratio | 2.40 [2.22, 3.05] | 2.60 [2.40, 4.20] | 0.305 |
| HDL | 55.50 [44.00, 68.25] | 63.00 [45.00, 64.50] | 0.961 |
| LDL | 72.00 [63.75, 86.00] | 84.00 [79.50, 85.50] | 0.222 |
| LDL.HDL.Ratio | 1.20 [1.10, 1.78] | 1.30 [1.20, 2.60] | 0.326 |
| Non.HDL.Cholesterol | 82.50 [74.75, 95.50] | 100.00 [96.00, 107.00] | 0.107 |
| Triglycerides | 54.50 [45.50, 87.00] | 87.00 [53.50, 133.00] | 0.157 |
| VLDL | 11.00 [9.00, 17.00] | 17.00 [11.00, 26.50] | 0.171 |
| Liverfat. | 0.55 [0.46, 0.62] | 0.49 [0.37, 5.04] | 1 |
| X.LiverFat..PDFF. | 1.17 [1.10, 1.59] | 1.25 [0.94, 4.96] | 0.77 |
| MRE.Average.Stiffness.in.kPa. | 1.85 [1.79, 2.15] | 1.94 [1.81, 2.17] | 0.596 |
| SAT_sum_Area.midfemur.slice | 18179.54 [13231.18, 21877.71] | 13069.77 [11578.63, 25190.13] | 0.558 |
| IMAT_sum_Area.midfemur.slice | 2059.82 [1268.76, 2771.35] | 1783.27 [1374.26, 3073.03] | 0.922 |
| Muscle_sum_Area.midfemur.slice | 26777.19 [23849.12, 29230.38] | 24363.55 [22914.31, 32462.90] | 0.922 |
| Bone_sum_Area.midfemur.slice | 1052.55 [908.29, 1171.28] | 979.34 [829.33, 1220.66] | 0.696 |
| SAT_sum_Volume | 1521280.91 [1113009.38, 1841803.88] | 1103042.32 [981185.76, 2087630.16] | 0.626 |
| IMAT_sum_Volume | 190781.10 [107734.79, 255509.14] | 170450.85 [127699.39, 274042.90] | 0.845 |
| Muscle_sum_Volume | 2151890.38 [1941567.85, 2364076.91] | 1950434.18 [1846165.17, 2626980.61] | 0.922 |
| Bone_sum_Volume | 87545.61 [75487.66, 96750.76] | 81232.92 [69983.96, 100696.56] | 0.696 |
| VAT.Mass_DEXA | 306.31 [255.11, 771.87] | 162.80 [123.46, 626.84] | 0.626 |
| VAT.Volume_DEXA | 324.69 [270.42, 818.18] | 172.56 [130.86, 664.45] | 0.626 |
| Total.Fat.Mass | 24440.52 [18851.99, 28248.50] | 21857.95 [18450.73, 33383.79] | 0.77 |
| Total.Lean.Mass | 43525.06 [40360.65, 53199.28] | 36569.48 [34871.68, 54471.06] | 0.435 |
| Total.Tissue.Mass | 71805.65 [60478.21, 76997.91] | 60760.70 [53492.07, 86003.96] | 0.77 |
| Total.Fat.Free.Mass | 46038.42 [42652.85, 55947.16] | 39285.42 [36944.93, 57863.11] | 0.435 |
| Total.Total.Mass | 74336.53 [62621.58, 80130.18] | 62939.71 [55801.90, 89396.02] | 0.77 |
| Arms.Region..Fat | 0.34 [0.25, 0.38] | 0.32 [0.27, 0.40] | 0.845 |
| Legs.Region..Fat | 0.36 [0.32, 0.43] | 0.37 [0.31, 0.41] | 0.845 |
| Trunk.Region..Fat | 0.35 [0.29, 0.38] | 0.34 [0.30, 0.43] | 0.558 |
| Android.Region..Fat | 0.36 [0.27, 0.41] | 0.36 [0.32, 0.46] | 0.626 |
| Gynoid.Region..Fat | 0.35 [0.33, 0.47] | 0.37 [0.31, 0.41] | 0.845 |
| Total.Region..Fat | 0.35 [0.28, 0.39] | 0.34 [0.29, 0.41] | 0.845 |
| Arms.Tissue..Fat | 0.35 [0.26, 0.39] | 0.33 [0.29, 0.41] | 0.845 |
| Legs.Tissue..Fat | 0.38 [0.33, 0.44] | 0.38 [0.32, 0.42] | 0.922 |
| Trunk.Tissue..Fat | 0.36 [0.29, 0.39] | 0.35 [0.31, 0.44] | 0.558 |
| Android.Tissue..Fat | 0.36 [0.27, 0.42] | 0.37 [0.32, 0.46] | 0.626 |
| Gynoid.Tissue..Fat | 0.36 [0.34, 0.48] | 0.38 [0.32, 0.42] | 0.77 |
| Total.Tissue..Fat | 0.36 [0.30, 0.40] | 0.36 [0.30, 0.42] | 0.77 |
| El..measured. | 2075.00 [1875.50, 2329.75] | 1765.00 [1656.00, 2497.00] | 0.626 |
| Nitrogen | 10.02 [8.86, 12.23] | 12.41 [9.31, 15.30] | 0.435 |
| X24Hour.EE | 2128.00 [1814.75, 2259.50] | 1767.00 [1594.00, 2531.50] | 0.77 |
| X24Hour.RQ | 0.89 [0.87, 0.91] | 0.88 [0.87, 0.89] | 0.66 |
| Exercise.EE..measured..kcal.min | 4.53 [3.98, 4.64] | 3.66 [3.50, 5.46] | 0.848 |
| Exercise.EE..measured..kcal.24.hr | 6516.00 [5735.00, 6685.00] | 5276.00 [5046.50, 7865.50] | 0.848 |
| ExerciseRQ | 0.92 [0.90, 0.97] | 0.92 [0.92, 0.93] | 0.949 |
| SleepEE | 1501.00 [1347.50, 1720.50] | 1255.00 [1151.50, 1805.00] | 0.696 |
| SleepRQ | 0.86 [0.84, 0.88] | 0.86 [0.84, 0.87] | 1 |
| NonExEE | 2028.00 [1782.50, 2068.00] | 1640.00 [1458.00, 2317.50] | 0.655 |
| NonExRQ | 0.87 [0.86, 0.88] | 0.88 [0.86, 0.88] | 0.565 |
| NonEx.NonSleep.EE | 2163.50 [1958.25, 2268.00] | 1804.00 [1617.50, 2527.00] | 0.495 |
| NonEx.NonSleep.RQ | 0.90 [0.87, 0.92] | 0.89 [0.88, 0.89] | 0.495 |
| EB | 39.00 [-7.00, 60.75] | 35.00 [-53.00, 62.00] | 0.922 |
| npRQ | 0.90 [0.88, 0.92] | 0.89 [0.88, 0.91] | 0.77 |
| ChoOx | 283.50 [246.75, 336.25] | 248.00 [205.50, 362.00] | 0.696 |
| FatOx | 61.00 [40.75, 77.25] | 59.00 [47.00, 66.50] | 0.626 |
| ProteinOx | 65.00 [57.00, 79.25] | 80.00 [60.50, 99.00] | 0.464 |
| Active.minutes | 153.50 [124.50, 173.75] | 120.00 [102.00, 130.50] | 0.079 |
| Activity. | 0.11 [0.09, 0.12] | 0.08 [0.07, 0.09] | 0.079 |
| SPA | 76.50 [60.75, 102.50] | 54.00 [32.50, 74.50] | 0.187 |
| TEF2..measured. | 0.06 [0.03, 0.07] | 0.04 [0.03, 0.06] | 0.922 |
| REE.pre.breakfast | 1609.00 [1528.25, 1851.25] | 1516.00 [1380.50, 1808.50] | 0.494 |
| RRQ.pre.breakfast | 0.87 [0.85, 0.87] | 0.86 [0.83, 0.86] | 0.283 |
| REE.post.breakfast | 1743.00 [1616.25, 1914.75] | 1571.00 [1440.00, 1977.50] | 0.495 |
| RRQ.post.breakfast | 0.92 [0.91, 0.94] | 0.90 [0.88, 0.92] | 0.143 |
| BEE | 1664.50 [1579.75, 1801.25] | 1391.00 [1357.00, 1989.50] | 0.495 |
| BRQ | 0.87 [0.86, 0.89] | 0.88 [0.87, 0.89] | 0.558 |
| Fat.Oxidation.Minutes..RQ...0.76. | 16.50 [4.25, 42.00] | 44.00 [4.50, 63.50] | 0.845 |
| Average.total.daily.bolus | 21.06 [15.96, 27.23] | NA [NA, NA] | NA |
| Average.bolus.dose.before.admission | 15.61 [14.87, 35.00] | NA [NA, NA] | NA |
| Average.bolus.dose.in.chamber | 25.74 [18.25, 26.00] | NA [NA, NA] | NA |
| Bolus.for.breakfast | 8.32 [4.31, 10.19] | NA [NA, NA] | NA |
| Daily.basal | 37.77 [25.65, 44.62] | NA [NA, NA] | NA |
| TTD | 60.28 [41.65, 75.43] | NA [NA, NA] | NA |
| TTD.in.chamber | 59.10 [49.55, 64.93] | NA [NA, NA] | NA |
| TTD.before.admission | 51.63 [37.34, 78.95] | NA [NA, NA] | NA |
| Pro..g..Consumed...breakfast | 17.10 [14.81, 18.25] | 15.29 [13.53, 18.45] | 0.435 |
| Fat..g..Consumed.breakfast | 12.12 [10.50, 13.15] | 11.21 [9.80, 13.90] | 0.558 |
| CHO..g..Consumed.breakfast | 118.90 [104.76, 128.73] | 108.87 [96.26, 131.70] | 0.626 |
| I.CHO.breakfast | 15.61 [12.30, 29.97] | NA [NA, NA] | NA |
| U.kg | 0.86 [0.74, 0.92] | NA [NA, NA] | NA |
| U.kg.in.chamber | 0.76 [0.72, 0.92] | NA [NA, NA] | NA |
| U.kg.before.admission | 0.80 [0.56, 0.96] | NA [NA, NA] | NA |
| bolus.kg.before.admission | 0.29 [0.23, 0.41] | NA [NA, NA] | NA |
| bolus.kg.total | 0.31 [0.25, 0.36] | NA [NA, NA] | NA |
| basal.kg | 0.48 [0.40, 0.56] | NA [NA, NA] | NA |
| Pancreas | 43322.21 [35921.24, 48231.46] | 78484.65 [63191.89, 94700.95] | 0.001 |
| Liver | 1437953.81 [1290598.69, 1725460.91] | 1185595.77 [1128007.52, 1661495.92] | 0.329 |
| Right.Kidney | 159300.90 [146297.12, 177952.06] | 140010.99 [118857.52, 166333.67] | 0.143 |
| Left.Kidney | 178556.03 [159871.62, 204226.50] | 122221.55 [118685.23, 173808.59] | 0.064 |
| Spleen | 263546.21 [230444.58, 320500.69] | 192669.65 [119813.74, 265899.14] | 0.143 |
| average.Pancreas.Fat.Fraction..exploratory. | 2.36 [1.93, 2.66] | 1.65 [1.50, 2.03] | 0.327 |
| MRE..kPa..where.applicable. | 0.59 [0.00, 1.33] | 0.00 [0.00, 1.94] | 0.788 |
| pancreas.tot_tiss_mass | 0.58 [0.50, 0.68] | 1.13 [0.99, 1.35] | 0.002 |
| Liver.tot_tiss_mass | 21.75 [19.71, 22.21] | 20.17 [19.32, 21.74] | 0.696 |
| Right_Kidn.tot_tiss_mass | 2.38 [2.05, 2.62] | 2.18 [1.81, 2.34] | 0.205 |
| left_Kidn.tot_tiss_mass | 2.58 [2.35, 2.93] | 2.15 [1.79, 2.48] | 0.097 |
| Spleen.tot_tiss_mass | 3.67 [3.22, 4.45] | 3.08 [2.26, 3.21] | 0.051 |
| pancreas.tot_tot_mass | 0.56 [0.48, 0.65] | 1.08 [0.96, 1.29] | 0.002 |
| Liver.tot_tot_mass | 20.95 [19.07, 21.33] | 19.60 [18.59, 20.79] | 0.558 |
| Right_Kidn.tot_tot_mass | 2.29 [1.99, 2.53] | 2.09 [1.75, 2.25] | 0.172 |
| left_Kidn.tot_tot_mass | 2.48 [2.28, 2.83] | 2.05 [1.73, 2.39] | 0.079 |
| Spleen.tot_tot_mass | 3.55 [3.11, 4.31] | 2.95 [2.17, 3.11] | 0.051 |
| pancreas.BW | 563.33 [490.38, 656.39] | 1100.96 [968.62, 1312.48] | 0.001 |
| Liver.BW | 21002.08 [19064.95, 21357.71] | 19815.41 [18790.20, 21159.16] | 0.558 |
| Right_Kidn.BW | 2273.41 [1965.35, 2593.80] | 2136.52 [1767.30, 2269.87] | 0.172 |
| left_Kidn.BW | 2486.42 [2286.47, 2904.74] | 2078.60 [1740.36, 2435.65] | 0.118 |
| Spleen.BW | 3592.92 [3196.23, 4247.35] | 3004.76 [2203.73, 3138.55] | 0.051 |

